# Associations of tumour somatic mutations with cancer-associated venous thromboembolism

**DOI:** 10.1101/2025.09.16.25335936

**Authors:** Naomi Cornish, Rebecca Ward, Matthew T. Warkentin, Chrissie Thirlwell, Andrew D. Mumford, Sarah K. Westbury, Philip C. Haycock

## Abstract

**Background:** Venous thromboembolism (VTE) is a common complication of cancer. Complex interactions between tumour biology and the haemostatic system may contribute to development of cancer-associated VTE.

**Objectives:** This study examined associations of somatic mutations with VTE in a large multi-cancer cohort.

**Methods:** We analysed paired tumour and germline whole genome sequence data and electronic health records from 12,507 cancer patients recruited to the Genomics England National Genomic Research Library, to evaluate associations of somatic mutations across 608 genes, overall tumour mutational burden (TMB) and 25 single base substitution (SBS) mutational signatures with VTE. Interactions between somatic mutations and a germline polygenic risk score for VTE were also assessed.

**Results:** In multivariable Cox regressions adjusted for age, sex and genetic ancestry, somatic mutations in four genes associated with higher rates of VTE at a false-discovery rate <0.1: *CDKN2A* (Hazard ratio, HR=1.62 [95% confidence interval, 1.23-2.13])*, KRAS* (HR=1.31 [1.12-1.53]), *PCDH15* (HR=1.48 [1.24-1.76]) and *TP53* (HR=1.55 [1.38-1.73]). SBS8, a common mutation signature of unknown aetiology, was also associated with higher rates of VTE (HR=1.39 [1.16-1.66]). In contrast, TMB ≥20 mutations/Mb, two DNA mismatch repair signatures (SBS6 and SBS26) and one rare signature of unknown aetiology (SBS19) associated with lower rates of VTE. Evidence for these associations remained robust after additional adjustment for tumour type, stage, and systemic anti-cancer treatment.

**Conclusions:** These findings support the hypothesis that tumour somatic mutations influence risk of VTE. This may provide insights into the pathophysiology of cancer-associated VTE and inform future efforts to improve clinical risk prediction.

## 1. Introduction

Venous thromboembolism (VTE) occurs in approximately 5-10% of patients with active cancer[1,2] and is a major cause of cancer related morbidity and mortality[3]. Identification of patients at highest risk of this condition is required to facilitate targeted interventions such as primary prophylaxis with anticoagulation[4].

Numerous clinical risk stratification tools for cancer-associated VTE have been developed over the past two decades[5]. The most extensively validated is the Khorana score, which stratifies patients according to primary tumour site, blood count parameters and body mass index[6]. However, the negative predictive value of the Khorana score is low, with >70% of VTE events occurring in patients who have not been categorized ‘high-risk’[7]. Efforts to improve risk prediction with more complex scoring algorithms which incorporate additional clinical variables have resulted in marginal improvements in performance[8].

Since VTE incidence varies significantly according to cancer-type[9], many risk scores apply a high weight to the primary tumour site[8]. This results in relatively poor risk stratification within cancer groups, particularly for cancers which are associated with high rates of VTE, including lung, pancreatic and gynaecological tumours[7].

Preclinical studies indicate that cancer cells may activate coagulation through a variety of intrinsic mechanisms, including tumour-induced platelet activation, over-expression of tumour-associated tissue-factor, or by inducing pro-inflammatory cytokine release and vascular endothelial damage[10–12]. Since somatic mutations alter tumour phenotype[13], exploring somatic genetic risks for VTE across many cancer types may help identify molecular determinants of cancer-associated thrombosis which apply to multiple anatomical tumour groups.

Three studies[14–16] have previously reported associations of somatic mutations with VTE in a pan-cancer (multiple cancer) setting. However, these studies were limited to targeted cancer-gene panels and many of the findings have not been replicated. To date, no studies have examined genome-wide somatic alterations and VTE.

The objective of this study was to evaluate associations of somatic mutations with VTE in a large pan-cancer cohort who had whole genome sequencing (WGS) performed on paired tumour and germline DNA. We assessed the effect of 1) tumour-somatic mutations in 608 protein-coding genes, 2) overall tumour mutational burden and 3) tumour mutational signatures on rates of VTE. As a secondary aim, we examined whether rates of VTE were influenced by interactions between a germline polygenic risk score (PRS) for VTE and somatic mutations within the tumour.

## 2. Methods

### 2.1 Cohort selection

The study cohort was derived from the 100,000 Genomes Project cancer programme, a Genomics England (GEL) initiative which prospectively recruited over 17,000 National Health Service (NHS) cancer patients of any age between 2015-2019 and performed whole genome sequencing (WGS) on matched germline and tumour DNA samples[17].

Genomic data is linked to pseudo-anonymised longitudinal electronic health records, including hospital episode statistics (HES), the National Cancer Registration and Analysis Service (NCRAS), the Systemic Anti-Cancer Therapy (SACT) dataset (which collates mandatory SACT reports from NHS providers), and mortality data from the Office for National Statistics (ONS).

From the recruited cohort (version 19 data-release), we selected any participant diagnosed with a solid organ or haematological malignancy who had WGS data meeting routine quality control thresholds (**Supplementary Methods**). Previously treated patients presenting with cancer recurrence, and patients recruited following neoadjuvant chemotherapy were included. We excluded patients where essential clinical information (including tumour-type or diagnosis date) was incomplete or conflicting between data sources; where tumour histology was subsequently reported as non-malignant; or where tumour WGS was performed on stored biopsies collected prior to study initiation. We also excluded patients with a history of VTE prior to study entry (defined as the date of tumour biopsy for WGS).

### 2.2 Outcome ascertainment

The primary outcome was first occurrence of VTE, defined as deep vein thrombosis at any anatomical site, or pulmonary embolism, identified from ICD10 codes[18] in hospital inpatient, outpatient and emergency department records (**Supplementary Table 1**).

Information on relevant covariates, including cancer type, stage and treatment was derived from information submitted by clinicians at GEL recruitment and the linked NCRAS and SACT data. Stage of cancer (recorded within 12months of tumour biopsy) was harmonized across different cancer types and categorized as early vs advanced (stage 3-4).

### 2.3 Genetic data

DNA extraction, WGS and variant calling were performed by GEL internal bioinformatic pipelines (described in **Supplementary Methods**). For participants with multiple tumour samples, we analysed WGS data from only one sample, prioritising in order of earliest collected sample; biopsy site (primary vs metastatic), tumour purity, preservation method (fresh frozen over formalin-fixed paraffin embedded), and WGS quality metrics respectively.

We filtered tumour WGS data to retain somatic single nucleotide variants (SNVs), small indels <50BP and large structural variants >50bp (SVs) that were predicted to impair canonical transcript function according to Cellbase (v.4.7.1) annotation[19], or predicted likely pathogenic according to COSMIC (v95)[20] or ClinVar (2018 release)[21], **Figure 1 and Supplementary methods**. For each participant sample, we applied a binary classification to every gene: mutated sequence if the participant carried one or more ‘filtered’ variants; otherwise, wildtype sequence.

**Figure 1:**
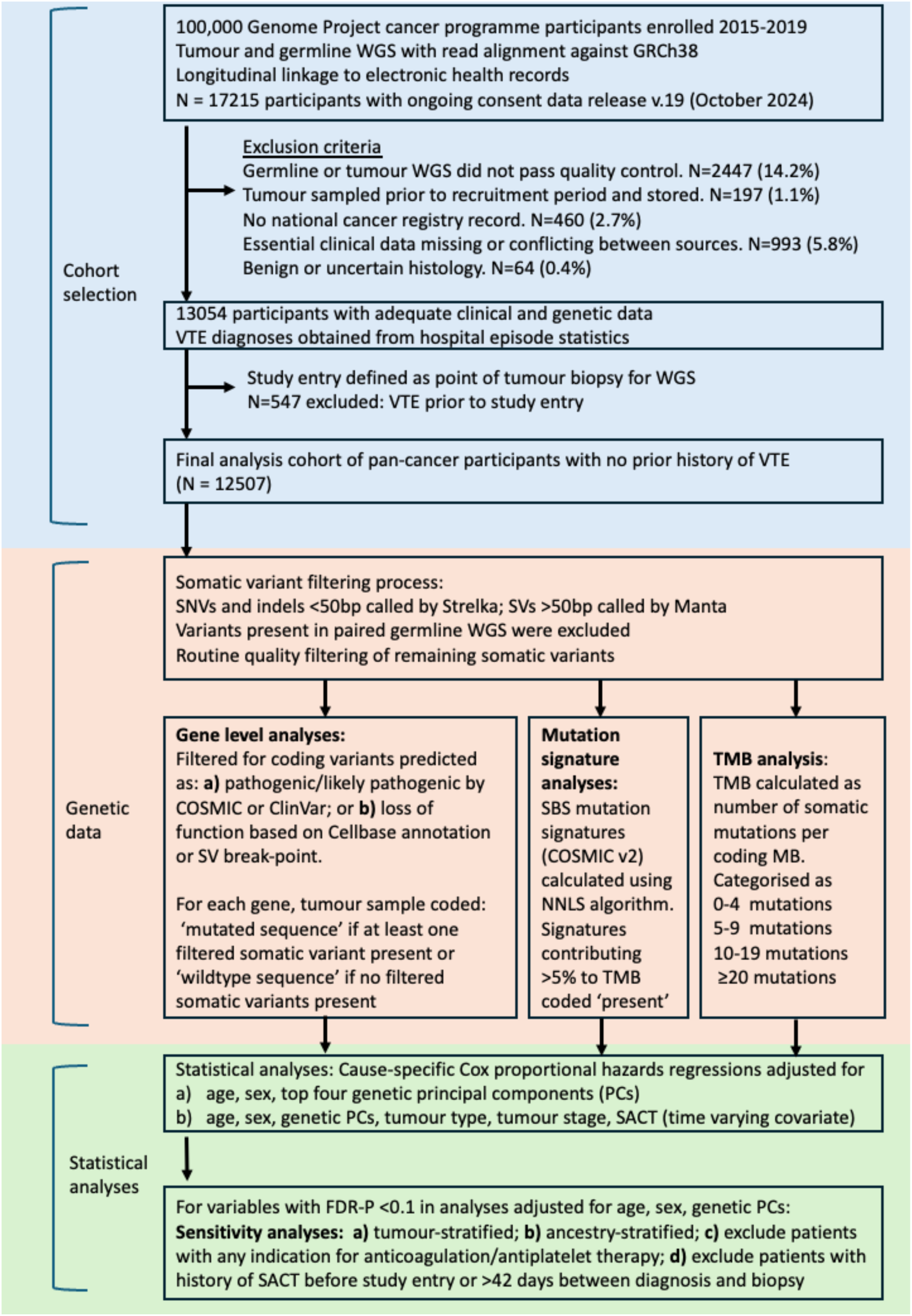
Flow chart showing study design. WGS, whole genome sequencing; VTE, venous thromboembolism; SNV, small nucleotide variant; SV, structural variant; SBS, single base substitution signature; NNLS, non-negative least squares; TMB, tumour mutational burden; FDR-P, false-discovery adjusted P value

In accordance with our published protocol[22], we analysed protein-coding genes (Ensembl v109)[23] where at least one of these criteria applied: 1) gene mutated in ≥5% of the cohort; 2) gene present in COSMIC cancer gene census v101[20] or 3) gene previously reported as a possible germline VTE locus in a published genome-wide association study (GWAS)[24–28]. To minimise the risk of unreliable results from sparse data[29] we excluded genes where fewer than 10 VTE events occurred in either the mutated or wildtype categories.

Tumour mutational burden (TMB) was calculated as the total number of somatic mutations per coding Mb in each sample and grouped into four categories: 0-4 mutations/Mb, 5-9 mutations/Mb, 10-19 mutations/Mb, and ≥20 mutations/Mb.

Single-base substitution (SBS) mutational signatures (defined by COSMIC Human Cancer Signatures v.2)[20], were calculated by GEL bioinformaticians using a non-negative least squares algorithm[30]. Signatures representing <5% of the overall mutation burden in a tumour sample were coded as zero values. Most signatures were present in only a subset of samples; therefore signatures were handled as a binary variable (present vs absent) and analyses restricted to comparisons with a minimum of 10 VTE events in each category.

For the VTE germline polygenic risk score (PRS), we used summary weights (log-odds ratios) from the ‘extended 297-SNP’ (single nucleotide polymorphism) PRS published by Klarin *et al*[24] which is derived from a VTE GWAS in a general population cohort. This PRS was chosen as it has previously been validated in a cancer population[31]. For each GEL participant (𝑗), the PRS was calculated from germline WGS data with PLINK version 2.0[32], using the default formula[33]: 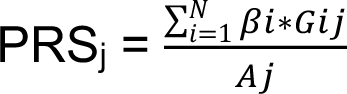 where 𝛽 is the risk-allele weight for each SNP (𝑖), 𝐺𝑖*j* is the risk allele count at SNP (𝑖) for participant (𝑗), 𝑁 is the number of VTE SNPs in the PRS and 𝐴𝑗 is the total number of valid allele observations for participant (𝑗). PRS scores were scaled to a mean of zero and standard deviation of 1. Genetic ancestry was inferred from principal components, calculated using germline SNPs as described in **Supplementary Methods.**

### 2.4 Statistical analyses

We evaluated the time from study-entry (date of tumour biopsy for WGS) to first episode of VTE, or death from any other cause. Participants were censored at the latest available date of electronic health record linkage (July 2022) or a maximum of 5 years from study entry.

We used cause-specific Cox proportional hazards (PH) regressions[34] to examine associations of somatic mutations (exposure) with VTE (outcome). Separate models were constructed to evaluate the effects of 1) somatic mutations in each gene; 2) TMB and 3) each SBS mutational signature on VTE.

Initial (‘minimally-adjusted’) analyses were adjusted for age, sex and the top four genetic principal components (PCs), which were considered likely confounders of the exposure-outcome relationship. P-values were adjusted for multiple testing using Benjamini & Hochberg false-discovery rate (FDR) correction. We then evaluated robustness of findings with additional ‘full’ adjustment for tumour type, stage, SACT during the study period, and history of SACT >6 weeks prior to study entry. These variables were hypothesised to correlate with both exposure and outcome, potentially acting as mediators of causal associations between somatic mutations and VTE (**Supplementary Figure 1**). SACT during the study period was evaluated as a time-varying covariate; patients who received SACT within 6 weeks prior to study entry were coded as exposed to SACT at baseline.

Tumour type was categorized according to GEL disease-groups used during recruitment (**Supplementary Table 2**). ‘Unavailable’ tumour-stage was included as a distinct category in the covariate adjustment, to avoid excluding participants with missing data for this variable.

For somatic mutation variables (genes, signatures, TMB) which were associated with VTE (FDR-P < 0.1 in minimally-adjusted analyses), we ran further sensitivity analyses to evaluate effect estimates after stratifying participants on the following criteria: 1) tumour type (to evaluate between-tumour heterogeneity); 2) genetically-inferred ancestry; 3) delay from cancer diagnosis to study entry (>42 days) or any history of SACT prior to study entry; and 4) any common medical indication for anticoagulation or antiplatelet therapy prior to study entry, including atrial fibrillation, ischaemic heart disease, or ischaemic stroke (**Supplementary Table 3**): these were used as proxy variables to exclude patients who were likely to be anticoagulated during the study, since prescription data were not available. We considered it unlikely that many patients would have received long-term anticoagulation as primary prevention for VTE; although some guidelines suggest consideration of VTE-prophylaxis in high-risk ambulatory cancer patients[35,36], routine implementation of these recommendations appears limited[37].

We also evaluated whether hazards remained proportional over the follow up period by examining time-dependent interaction terms and time-stratified analyses. We assessed the cumulative incidence for VTE and sub-hazard ratio for VTE associated with each variable after accounting for death as a competing risk factor with Fine and Gray regression models. Finally, we evaluated the interactions between somatic mutations and a germline VTE-PRS.

A peer-reviewed a-priori protocol for this analysis is published in Wellcome open research[22]. There were minimal deviations from the protocol, these are outlined in **Supplementary Methods.**

## 3. Results

### 3.1 Study cohort

Of the 17,215 participants originally recruited to the 100,000 Genomes Project cancer programme, we excluded 4161 participants due to inadequate quality WGS or missing essential clinical data **(Figure 1).** A further 547 participants were excluded as they had a VTE diagnosis before study entry; 380 of these excluded VTE events occurred before cancer diagnosis, while 167 VTE events occurred between cancer diagnosis and study entry. In total, 12,507 participants were included in the analyses. Median time from diagnosis to study entry (tumour biopsy for WGS) was 42 days (interquartile range; IQR, 18-94 days) and median follow up for the cohort was 50 months (IQR, 36-59 months).

Over the observation period, 1,177 participants experienced a VTE and 2,816 participants died without a diagnosis of VTE. The initial rate of VTE declined steeply over the first 12 months, then plateaued **(Supplementary Figure 2**). Cohort characteristics are shown in **Table 1**. In univariable Cox PH regression models, clinical factors which associated with higher rates of VTE (p<0.05) included African ancestry (compared with European ancestry), advanced stage cancer, SACT and tumour type (namely adult glioma, hepatopancreatobiliary, lung, ovarian, and upper gastrointestinal tumours, when comparing each tumour type to all others), **Table 2**.

**Table 1.** Cohort characteristics stratified by VTE status at end of follow up

|  | Status at end of follow up |  | Overall, n = 12507 |
| --- | --- | --- | --- |
|  | No VTE , n = 11330 | VTE, n = 1177 |  |
| Sex |  |  |  |
| Female | 6531 (58%) | 660 (56%) | 7191 (58%) |
| Male | 4799 (42%) | 517 (44%) | 5316 (42%) |
| Age (years) |  |  |  |
| Median (interquartile range) | 65 (55-73) | 65 (55-73) | 65 (55-73) |
| Genetically inferred ancestry |  |  |  |
| European | 10135 (90%) | 1047 (89%) | 11182 (89%) |
| African | 419 (4%) | 62 (5%) | 481 (4%) |
| American | 218 (2%) | 15 (1%) | 233 (2%) |
| East Asian | 113 (1%) | 5 (0%) | 118 (1%) |
| South Asian | 445 (4%) | 48 (4%) | 493 (4%) |

| **Cancer type** |  |  |  |
| --- | --- | --- | --- |
| Breast | 2495 (22%) | 202 (17%) | 2697 (22%) |
| Colorectal | 2164 (19%) | 222 (19%) | 2386 (19%) |
| Lung | 1149 (10%) | 155 (13%) | 1304 (10%) |
| Renal | 1107 (10%) | 98 (8%) | 1205 (10%) |
| Sarcoma | 711 (6%) | 91 (8%) | 802 (6%) |
| Endometrial carcinoma | 670 (6%) | 86 (7%) | 756 (6%) |
| Ovarian | 449 (4%) | 71 (6%) | 520 (4%) |
| Prostate | 493 (4%) | 21 (2%) | 514 (4%) |
| Haematological | 453 (4%) | 30 (2%) | 483 (4%) |
| Adult glioma | 414 (4%) | 57 (5%) | 471 (4%) |
| Bladder | 312 (3%) | 32 (3%) | 344 (3%) |
| Hepatopancreatobiliary | 223 (2%) | 44 (4%) | 267 (2%) |
| Malignant melanoma | 219 (2%) | 26 (2%) | 245 (2%) |
| Head and neck | 214 (2%) | 13 (1%) | 227 (2%) |
| Upper gastrointestinal<sup>1</sup> | <221 | <30 | 221 (2%) |
| Other<sup>1</sup> | <65 | <5 | 65 (<1%) |
| **Stage of tumour** |  |  |  |
| early | 5977 (53%) | 485 (41%) | 6462 (52%) |
| advanced | 3026 (27%) | 456 (39%) | 3482 (28%) |
| unknown | 2327 (20%) | 236 (20%) | 2563 (20%) |
| **Treatment** |  |  |  |
| History of SACT<sup>2</sup>>6weeks prior to study | 1746 (15%) | 265 (22%) | 2011 (16%) |
| SACT during study period | 4540 (40%) | 662 (56%) | 5202 (42%) |
| Surgery during study period<sup>3</sup> | 10517 (93%) | 1110 (94%) | 11627 (93%) |

<sup>1</sup>exact counts for 'upper gastrointestinal' and 'other' are masked to prevent derivation of counts < 5, as per GEL data governance rules; <sup>2</sup>SACT, systemic anti-cancer therapy; <sup>3</sup>Surgery occurring within 6 weeks prior to study entry or any time following study entry.

**Table 2:**
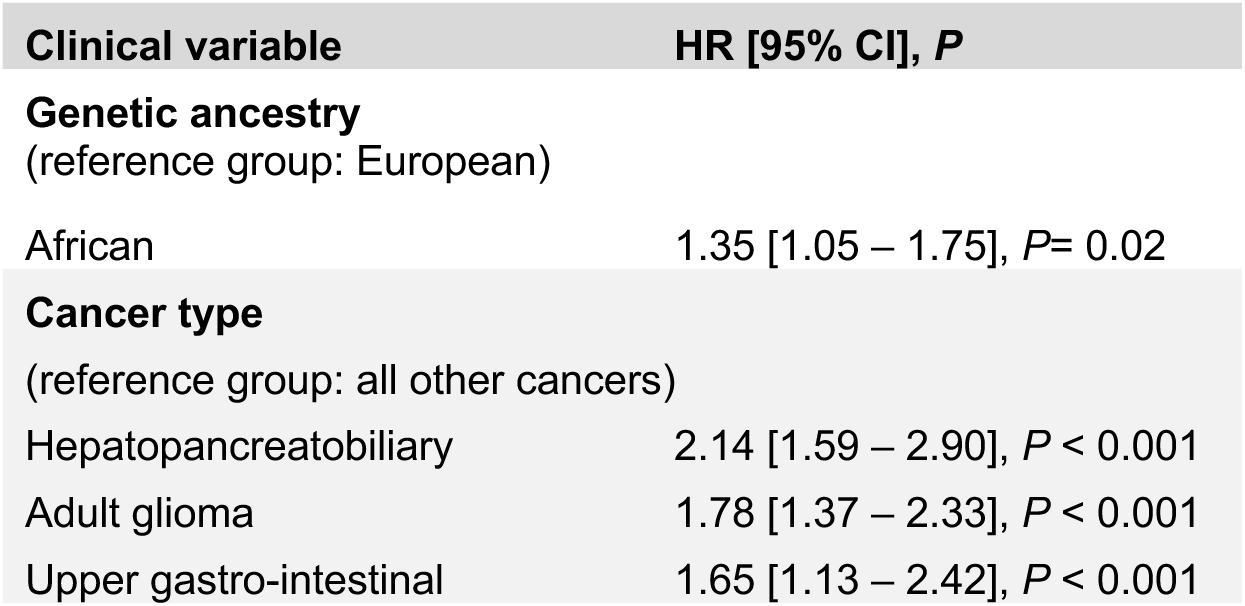
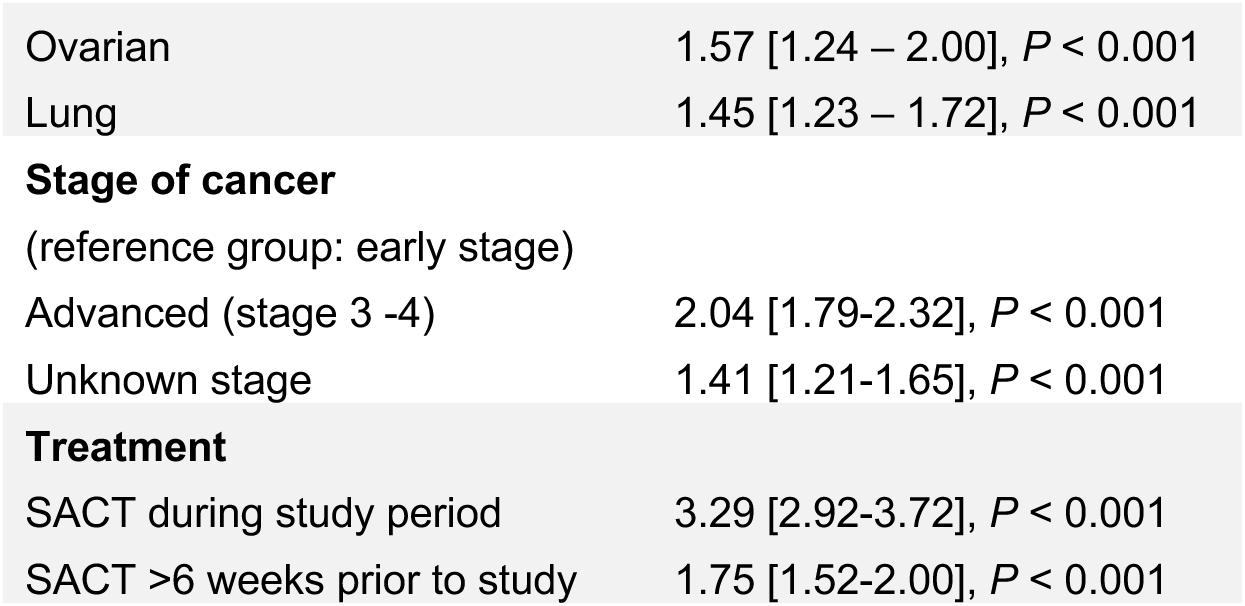
Associations of clinical variables with VTE assessed using univariable Cox PH regressions. HR, hazard ratio; CI, confidence interval; P, p-value. Only variables associated with increased rate of VTE (p<0.05) are shown. Full results for all variables are in Supplementary Table 4.

| **Clinical variable** | **HR [95% CI], P** |
| --- | --- |
| **Genetic ancestry**<br>(reference group: European) |  |
| African | 1.35 [1.05 – 1.75], $P = 0.02$ |
| **Cancer type**<br>(reference group: all other cancers) |  |
| Hepatopancreatobiliary | 2.14 [1.59 – 2.90], $P < 0.001$ |
| Adult glioma | 1.78 [1.37 – 2.33], $P < 0.001$ |
| Upper gastro-intestinal | 1.65 [1.13 – 2.42], $P < 0.001$ |
| Ovarian | 1.57 [1.24 – 2.00], $P < 0.001$ |
| Lung | 1.45 [1.23 – 1.72], $P < 0.001$ |
| Advanced (stage 3 -4) | 2.04 [1.79-2.32], $P < 0.001$ |
| Unknown stage | 1.41 [1.21-1.65], $P < 0.001$ |
| Treatment |  |
| SACT during study period | 3.29 [2.92-3.72], $P < 0.001$ |
| SACT >6 weeks prior to study | 1.75 [1.52-2.00], $P < 0.001$ |

### 3.2 Somatic mutations in protein-coding genes and VTE

We examined effects of somatic mutations across 608 genes on rates of VTE. Initial Cox PH regressions, referred to as minimally adjusted models, were adjusted for age, sex and genetic PCs. Applying an FDR <0.1, we found evidence that four genes were associated with higher rates of VTE: *TP53* (Hazard ratio, HR=1.55 [95% confidence interval, 1.38-1.73], *P*=1.25×10^-13^, FDR-P<0.001), *PCDH15* (HR=1.48 [1.24-1.76], *P*=9.44×10^-6^, FDR-P=0.003), *CDKN2A* (HR=1.62 [1.23-2.13], *P*=5.6×10^-4^, FDR-P=0.094); and *KRAS* (HR=1.31 [1.12-1.53], *P*=6.21×10^-4^, FDR-P=0.094). Pan-cancer effect estimates for *CDKN2A*, *KRAS* and *PCDH15* were similar in Cox PH regressions where we additionally included tumour site, stage and SACT as covariates, whereas for TP53 the association with VTE was somewhat attenuated **(Figure 2, Supplementary Table 5)**.

**Figure 2:**
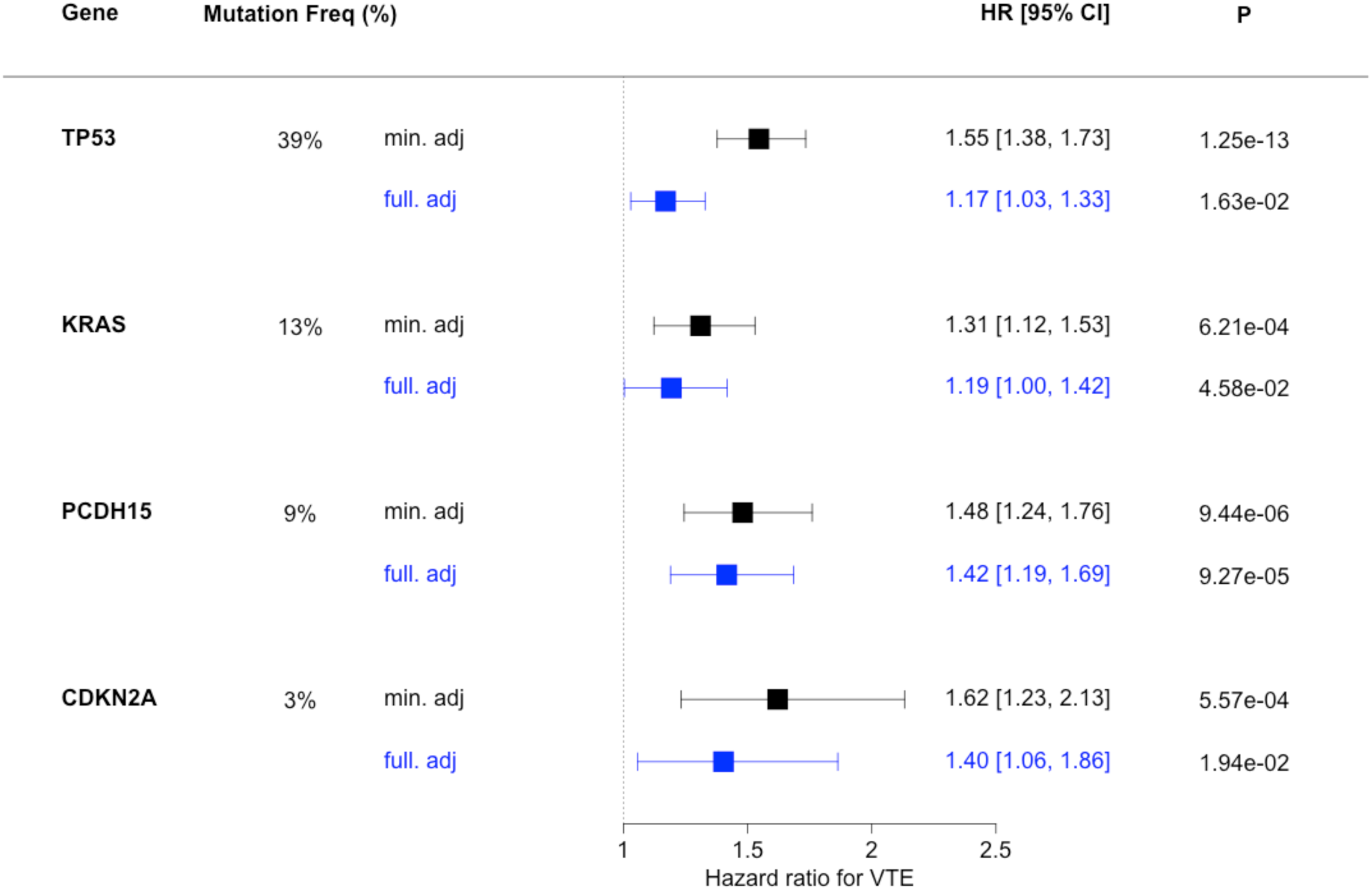
Hazard ratios (HR), 95% confidence intervals (CI) and unadjusted p-values (P) for pan-cancer associations of gene-level somatic mutations with VTE (associations with FDR-P < 0.1 in minimally adjusted analyses are shown). Black plots (min. adj) show analyses adjusted for age, sex, top four genetic PCs. Blue plots (full. adj) show analyses adjusted for age, sex, top four genetic PCs, tumour type, tumour stage, history of SACT more than 6 weeks before study entry and SACT during study period (time-varying covariate). Mutation Freq (%), indicates the percentage of tumour samples with at least one somatic mutation in the gene.

Inclusion of time-dependent interaction terms and time-stratified analyses **(Supplementary Table 6)** indicated that associations of mutations in *CDKN2A*, *KRAS* and *PCDH15* with VTE remained relatively constant over time. For *TP53* there was a significant reduction in the association with VTE over time; HR 1.74 [1.47-2.07] in year 1 compared to HR 0.93 [0.58-1.51]) in year 5; (TP53:year5 time interaction term *P*=0.02). Effect estimates for all four genes were similar after accounting for death as a competing risk factor in Fine and Gray risk regressions (**Supplementary Table 6**) and somatic mutations in these genes were associated with a higher estimated cumulative incidence of VTE **(Supplementary Figure 3).**

Tumour-stratified analyses indicated that, for many tumour types, the direction of effect for each gene-VTE association appeared consistent with the pan-cancer signal. However, confidence intervals for most tumours were wide and included the possibility of no effect; for some tumours, regression models did not converge due to insufficient numbers in each category. Point estimates for gene-VTE associations were occasionally in the opposite direction to the pan-cancer results, for example: malignant melanoma (*TP53, PCDH15* and *CDKN2A*); haematological cancers (*TP53* and *KRAS*) and adult glioma (*TP53* only) **(Figure 3, Supplementary Table 6)**. Overall, we did not see strong evidence for between-tumour heterogeneity in the estimates for each gene-VTE association as measured by Cochran’s Q statistics (**Supplementary Table 7)**.

**Figure 3:**
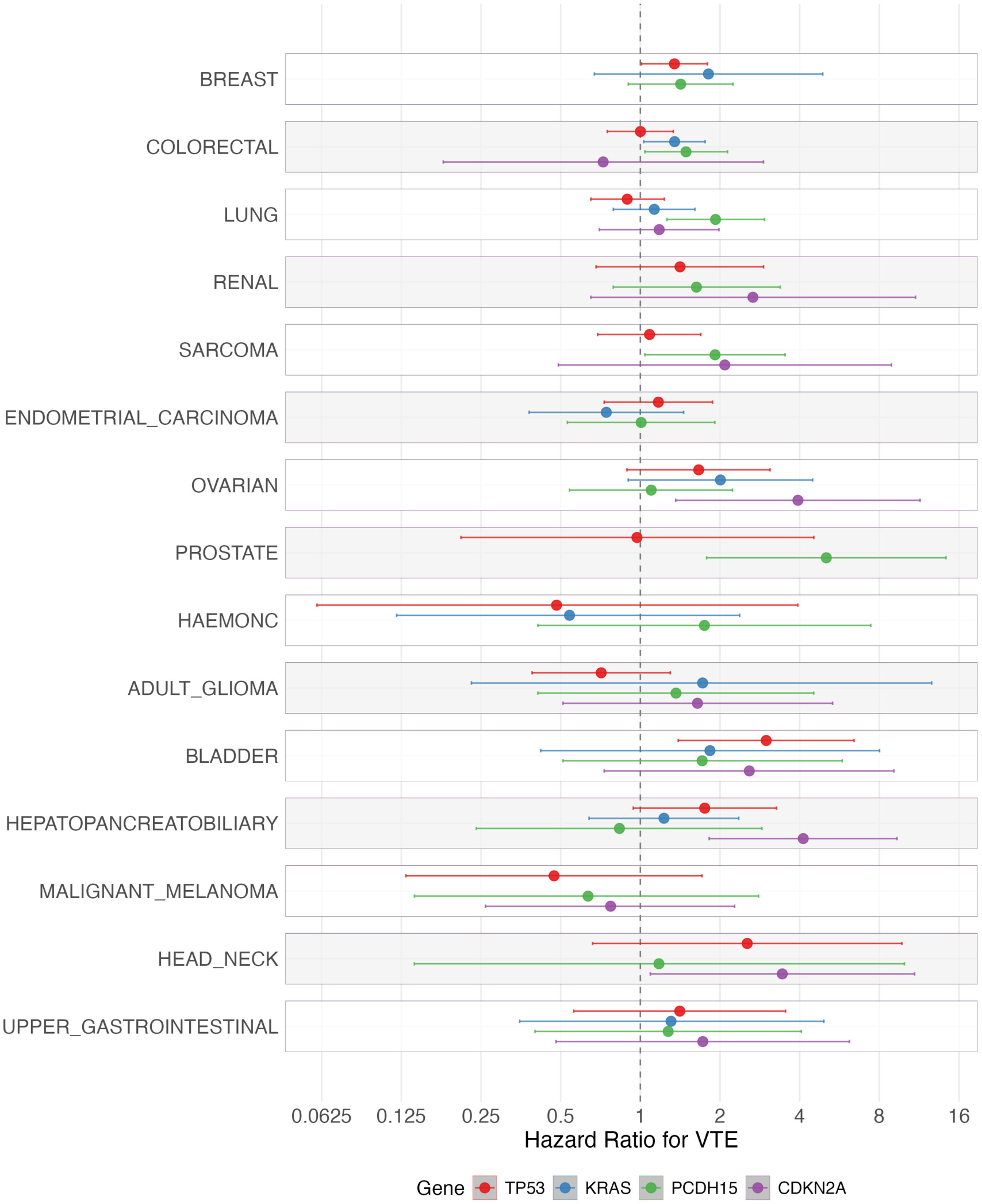
Associations of somatic mutations in TP53, KRAS, PCDH15 and CDKN2A with VTE, stratified by tumour type. Results from Cox PH models (adjusted for age, sex, genetic PCs, tumour stage and SACT). If no estimate is shown, the data was too sparse to allow stable regression model estimation within that cancer type. The x-axis has been plotted on a log2 scale.

Pan-cancer VTE effect estimates for all four genes remained relatively consistent in sensitivity analyses where we 1) excluded patients with a medical indication for either anticoagulation or anti-platelet therapy 2) restricted analyses to patients who entered the study within 42 days of initial cancer diagnosis and had not received prior SACT and 3) included *TP53, KRAS, PCDH15* and *CDKN2A* in a multi-gene regression model. Ancestry-stratified estimates for non-Europeans were imprecise due to small sample sizes but generally trended in the same direction as the main results **(Supplementary Table 6)**.

### 3.3 Tumour mutational burden, SBS mutational signatures and VTE

We analysed associations of TMB and 25 SBS mutational signatures with VTE. There was a non-linear association between TMB and VTE: TMB≥20 mutations/Mb was inversely associated with VTE (HR 0.60 [0.45-0.80], *P*=5×10^-4^; fully-adjusted model) when compared to the lowest TMB category (0-4 mutations/Mb). In contrast, TMB between 5-9 or 10-19 mutations/Mb was not associated with VTE after full adjustment for tumour type, stage and SACT (**Table 3**). Effect estimates for the association between TMB≥20 mutations/Mb and VTE were similar when TMB was categorized as a binary variable (comparing TMB≥20 with TMB<20 mutations/Mb), **Figure 4, Supplementary Table 8**.

**Table 3.** Results from Cox PH regressions showing associations of 1) tumour mutational burden (TMB) with VTE and 2) COSMIC v2 single base substitution (SBS) mutational signatures with VTE. HR, Hazard ratio; 95% CI, 95% confidence interval; TMB, tumour mutational burden; SBS, single base substitution signature. Minimally adjusted models were adjusted for age, sex, genetic principal components; fully adjusted models included additional adjustment for tumour type, stage and systemic anti-cancer therapy (SACT). Variables which associated with VTE at an FDR-P < 0.1 in the minimally-adjusted models are shown. *Associations which persisted in fully-adjusted analyses (P<0.05). Full results for all signatures are in Supplementary Table 9.

|  | Minimally adjusted model | Fully adjusted model |
| --- | --- | --- |
|  | HR[95% CI], P | HR[95% CI], P |
| **TMB category**<br>(reference category: 0-4 mutations/Mb) |  |  |
| 5-9 coding mutations / Mb | 1.34 [1.12-1.60], P=0.001 | 1.10 [0.91-1.34], P=0.308 |
| 10-19 coding mutations / Mb | 1.34 [1.02-1.74], P=0.033 | 1.07 [0.80-1.43], P=0.651 |
| ≥20 coding mutations / Mb* | 0.62 [0.47-0.81], P=0.001 | 0.60 [0.45-0.80], P<0.001 |
| **SBS mutational signatures**<br>(reference category: signature absent) |  |  |
| SBS4 (Tobacco smoking) | 1.29 [1.08-1.53], P=0.004 | 1.04 [0.80-1.35], P=0.757 |
| SBS5 (Unknown aetiology) | 0.81 [0.72-0.91], P=0.000 | 0.96 [0.84-1.10], P=0.555 |
| SBS6 (Defective DNA mismatch repair)* | 0.64 [0.48-0.86], P=0.003 | 0.73 [0.54-0.99], P=0.041 |
| SBS8 (Unknown aetiology)* | 1.39 [1.16-1.66], P=0.000 | 1.36 [1.12-1.65], P=0.002 |
| SBS17 (Unknown aetiology) | 1.35 [1.08-1.68], P=0.007 | 1.09 [0.85-1.39], P=0.512 |
| SBS19 (Unknown aetiology)* | 0.58 [0.42-0.80], P=0.001 | 0.63 [0.45-0.89], P=0.008 |
| SBS26 (Defective DNA mismatch repair)* | 0.54 [0.32-0.91], P=0.021 | 0.50 [0.29-0.86], P=0.012 |
| SBS30 (Defective base excision repair due to NTHL1 inactivation) | 0.79 [0.63-0.98], P=0.032 | 0.88 [0.70-1.10], P=0.255 |

**Figure 4.**
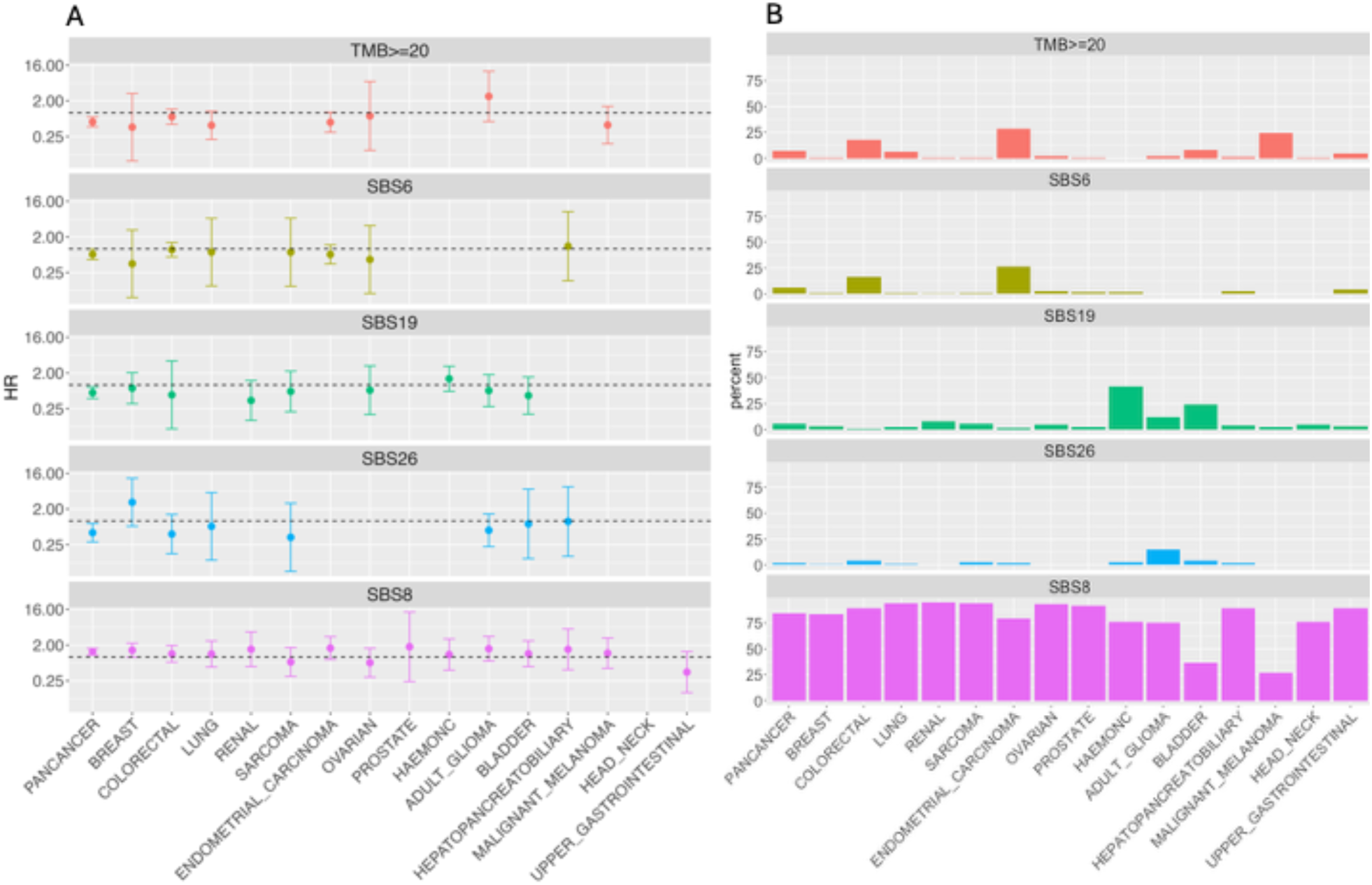
A (LEFT) Pan-cancer and tumour stratified analyses showing associations of TMB≥20 (compared with TMB<20) and mutational signatures SBS6, SBS19, SBS26 and SBS8 with VTE. Results (Hazard ratio, HR, and 95% confidence intervals) from Cox models with adjustment for age, sex, genetic PCs, tumour stage, SACT are shown. Tumour type is ranked in order of overall sample size. If no estimate is shown, the data was too sparse for stable regression model estimation. The y-axis is plotted on a log2 scale. HR, hazard ratio for VTE. B (RIGHT) Prevalence (%) of TMB≥20 and mutation signatures SBS6, SBS19, SBS26 and SBS8 within each cancer type.

We similarly observed that DNA mismatch repair signatures SBS6 and SBS26 were associated with lower rates of VTE in both minimally adjusted analyses (FDR-P<0.1) and fully adjusted analyses (SBS6, HR=0.73 [0.54-0.99]; SBS26, HR=0.50 [0.29-0.86]). SBS19 (a signature of unknown aetiology) was also associated with lower rates of VTE (HR=0.63 [0.45-0.89]), while SBS8 (unknown aetiology) was associated with higher rates of VTE (HR=1.36[1.12-1.65]), **Table 3, Supplementary Table 9**.

There were no significant changes to the pan-cancer results for TMB or mutational signatures across the various pre-planned sensitivity analyses (described in methods); results were also consistent in a further sensitivity analysis where we adjusted for treatment with immune check-point inhibitors (**Supplementary tables 8-10**).

However, in a sensitivity analysis where each signature was examined as a continuous variable (restricted to samples where the signature was present), we did not find evidence for a dose-dependent relationship between any signature and VTE (i.e. no association between the total contribution of the signature to mutation burden and VTE), **Supplementary Table 10**.

Since TMB and the presence of specific mutational signatures is highly dependent on tumour type[38], we evaluated the distribution of TMB and signatures within each tumour group to inform our interpretation of results (**Figure 4**, **Supplementary Tables 8-10).** High TMB (≥20 mutations/Mb) was relatively frequent in colorectal cancer (18%), endometrial cancer (29%) and malignant melanoma (25%). In tumour-stratified analyses, high TMB trended towards reduced VTE risk across those tumour types (endometrial cancer, HR 0.57 [0.32-1.03]; malignant melanoma, HR 0.49 [0.16-1.45]; colorectal cancer, HR 0.79 [0.51-1.23]), as well as for lung cancer (HR 0.48 [0.21-1.09]) and possibly breast cancer, where only a small proportion of tumours (1%) had TMB≥20, reflected by wide confidence intervals for the effect estimate (HR 0.43 [0.06-3.09]). For many cancers it was not possible to run stratified analyses as there were insufficient VTE events in the TMB≥20 category. Sequentially excluding cancer types with a high median TMB in leave-one out analyses did not alter the overall pan-cancer result **(Supplementary Figures 4-5)**.

SBS8 was common across all tumour types (identified in 85% of all tumours) and on average comprised just under 20% of the total mutational burden of each tumour (**Supplementary Figure 6**). In contrast the other signatures were relatively rare, except in specific cancers: SBS6 was found in >15% of colorectal cancer and endometrial cancer samples, SBS26 was seen most frequently in glioma (15% of samples) and SBS19 was predominantly seen in haematological and bladder cancers (41% and 25% respectively). In all cases, tumour-stratified estimates for associations of each signature with VTE showed very wide confidence intervals making it difficult to draw any conclusions, however, effect estimates appeared broadly consistent with the pan-cancer signal, with the notable exception of SBS19 in haematological cancers (HR for VTE 1.43 [0.69-2.97]).

### 3.4 Interactions between germline polygenic risk score (PRS), somatic mutations and VTE

A germline VTE PRS-score was calculated for each participant using SNPs and weights derived from a VTE GWAS by Klarin *et al*[24]. We excluded 120 patients from the PRS analyses as their germline WGS data was not included in the quality-filtered aggregate genotype files for the cohort. Cox PH regressions minimally adjusted for age, sex and genetic PCs showed the rate of VTE increased (HR = 1.18 [1.12-1.25], P = 9.56x 10^-10^) for every standard deviation rise in PRS. We also observed an interaction between the germline PRS score and *PCDH15* (*P*=0.003 for the PRS:PCDH15 interaction term); the association of somatic mutations in *PCDH15* with VTE appeared greater in people with a high germline PRS (top quartile) compared to people with a lower germline PRS (lower three quartiles) (**Figure 5**; **Supplementary Table 11**). There was no robust evidence for interactions between the PRS and somatic mutations in *TP53, KRAS* or *CDKN2A*, nor for interactions between the PRS and TMB or SBS mutational signatures.

**Figure 5:**
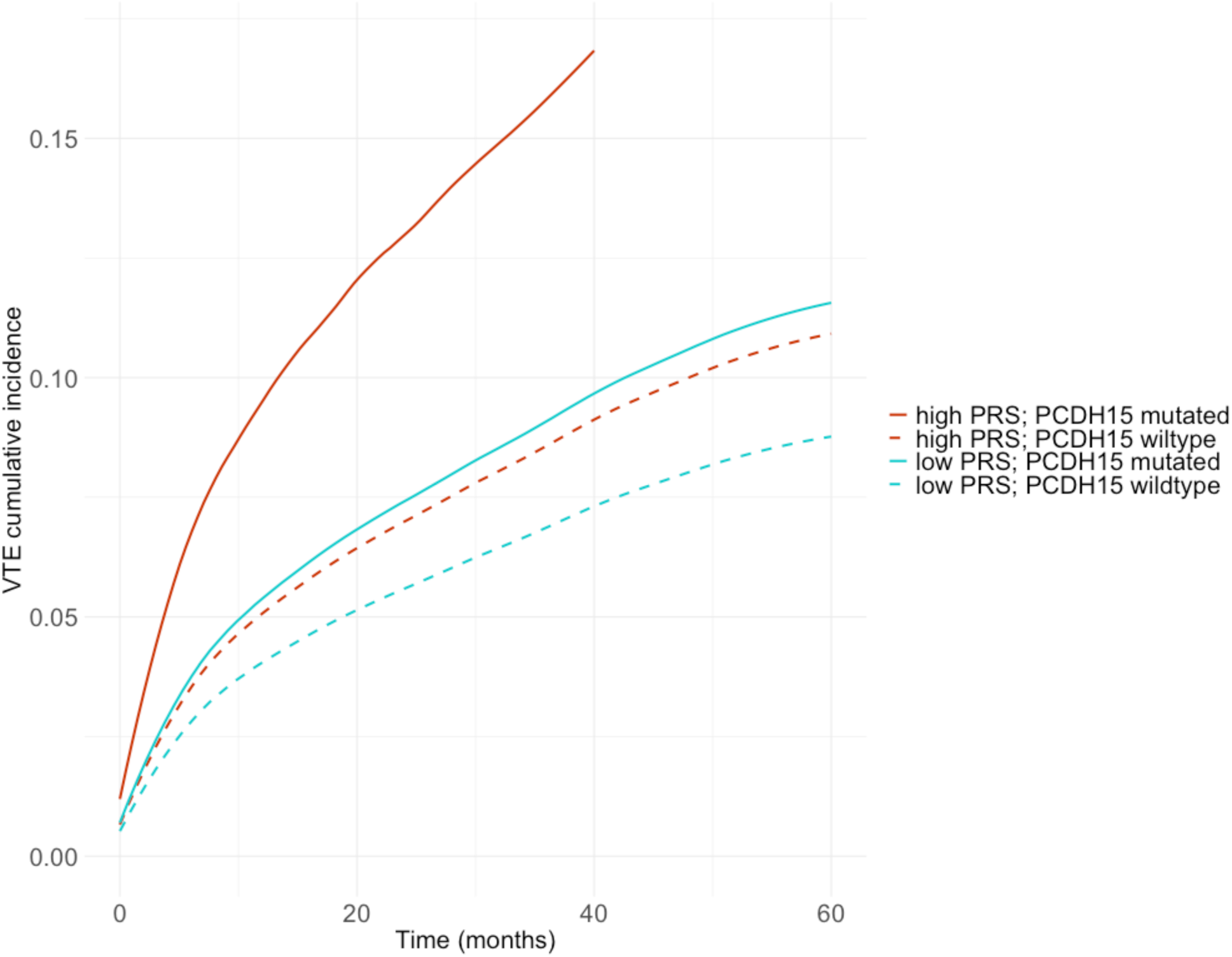
Cumulative incidence for VTE after stratifying on both germline PRS and somatic PCDH15 mutation status. Red (high PRS); Blue (low PRS), solid lines (PCDH15 mutated); dotted lines (PCDH15 wildtype).

## 4. Discussion

Characterizing tumour-genetic risks for VTE may provide insights into the pathophysiology of cancer-associated VTE and inform efforts to improve clinical risk prediction. This study examined associations of somatic mutations in 608 genes, TMB, and 25 SBS mutational signatures with VTE. We found higher rates of VTE associated with somatic mutations in four genes (*CDKN2A, KRAS, PCDH15* and *TP53*) and SBS8. We found lower rates of VTE in the context of TMB≥20 mutations/Mb, two signatures associated with DNA mis-match repair[39] (SBS6 and SBS26) and one signature of unknown aetiology (SBS19).

Associations persisted after adjusting for tumour-type, stage and SACT in ‘fully adjusted’ multivariable Cox-models, and across numerous sensitivity analyses, although the effect estimate for the association between *TP53* and VTE attenuated to some degree in the fully adjusted pan-cancer model, and at in the final year of observation in the time-stratified analyses.

Somatic mutations in *CDKN2A*, *KRAS* and *TP53* have all been implicated in other observational studies of cancer-associated thrombosis [14,15,40], therefore our results serve as useful independent replication of previous reports. Associations of *PCDH15*, SBS mutational signatures and TMB with VTE have not been described previously.

This is an observational analysis of real-world data, therefore causal inference is challenging and should be approached with caution. One hypothesis is that somatic mutations contribute directly to the pathophysiology of VTE, for example by inducing tumour-expression of pro-thrombotic proteins. There is supportive evidence of this for *KRAS*, where experiments in colorectal cancer cell-lines have shown that activating mutations in the gene result in upregulation of tissue-factor expression[41]*. TP53* deficiency (and pharmaco-inhibition of the p53 protein) in mice has also been associated with increased levels of circulating tissue factor and impaired VTE resolution[41,42].

*CDKN2A* encodes a protein (p16^INK4A^) which inhibits cyclin-dependent kinases CDK4 and CDK6[43]. Mutations in *CDKN2A* may potentially disrupt a range of biological pathways, including CDK6-NFkB mediated inflammatory chemokine release[44], or CDK6-regulated neutrophil-extracellular trap formation[45], which could potentially contribute to thrombosis. *CDKN2A* loss in murine-models leads to abnormal proliferation of platelets[46] and pro-inflammatory macrophages[47], and germline SNPs near the *CDKN2A* locus on chromosome 9p21.3 have been associated with arterial thromboses in genome-wide association studies of the general population[48]. This evidence supports the hypothesis that *CDKN2A* may be causally implicated in the development of cancer-associated VTE.

However, *CDKN2A, TP53 and KRAS* are genes with a known role in tumorigenesis and influence cell-cycle regulation or cell-signalling respectively[20]. Mutations in these genes may therefore proxy more aggressive histological subtypes of malignancy, leading to indirect associations with VTE. For example; larger, more advanced tumours may compress or invade vessels, contributing to venous stasis and subsequent thrombosis[49].

Patients with advanced disease are also more likely to require SACT, have prolonged periods of hospitalization or experience complications such as infection, all of which are associated with VTE[49,50]. In the era of molecularly-targeted therapies, selection of treatments may also be dictated by knowledge of somatic mutation profile[13], resulting in associations between somatic mutations and VTE which are driven through the action of drugs, rather than any direct biological phenomenon.

In the tumour-stratified analyses, we found directions of effect for the associations of *CDKN2A, TP53 and KRAS* with VTE appeared broadly consistent across most (but not all) tumour types, albeit with wide variations in estimated effect sizes **(Figure 3, Supplementary Table 6)**. This may be attributable to a multitude of complex factors which mediate risks of VTE, including variations in mutation frequencies, gene-gene interactions, differences in tumour micro-environment or specific treatment-related interventions (e.g. central venous catheter insertion or chemotherapeutic combinations). Further tumour-specific analyses in large cohorts, in conjunction with mechanistic studies in pre-clinical models, may help understand the context in which these genes increase thrombotic risk.

We observed a novel association between *PCDH15* and VTE. This gene has not been investigated in previous studies of cancer-associated VTE. *PCDH15 is* a member of the cadherin superfamily, which encode integral membrane proteins involved in calcium-dependent cell adhesion. Germline variants in this gene are associated with Usher syndrome, a hereditary disorder of hearing and vision which is not known to be associated with thrombosis[51]. Somatic mutations in *PCDH15* have recently been associated with metastatic disease in prostate cancer[52,53] and ocular carcinoma[53]. Evidence from *in vitro* functional studies suggests it may be involved in tumour cell proliferation through interactions with the WNT/b-catenin signalling pathway[54]. Interestingly, in tumour-stratified analyses, *PCDH15* showed the strongest association with VTE in prostate cancer (HR=5.04 [1.78-14.28], *P*=0.002) and smaller but significant (*P*<0.05) associations in sub-group analyses for sarcoma, lung and colorectal cancer.

The association between somatic *PCDH15* mutations and VTE was also greater in participants with a high germline PRS for VTE, compared to those with a lower germline PRS **(Figure 5)** Since tumours that create a pro-coagulant niche may have greater metastatic potential[10,55], one possibility is that *PCDH15* mutations contribute to reciprocal interactions between tumour progression and the host haemostatic system.

However, 75% of the somatic *PCDH15* variants in our cohort were structural variants (SVs) >50bp, mainly non-fusion translocations. SVs involving the *PCDH15* locus could potentially disrupt distal cis-or trans-regulatory elements for other genes, making it unclear whether the observed effect on VTE is specific to *PCDH15*. Since SV calls from short-read sequence data have limited precision and may include a high proportion of false positives[56], replication of this finding in independent cohorts, ideally using long-read WGS, would be valuable before drawing conclusions.

Endogenous biological processes and exogenous mutagenic exposures may shape the pattern of somatic mutations in a tumour, leading to recognizable mutational signatures within the cancer genome[30]. To our knowledge this is the first study to report on COSMIC SBS mutational signatures and risk of VTE. We observed an inverse association between DNA mismatch repair signatures[39] SBS6 and SBS26, as well as high TMB ≥20 mutations/Mb, and VTE.

DNA mismatch repair deficiency correlates with high TMB[57], which in turn has been shown to be associated with better progression-free and overall survival in patients treated with immune checkpoint inhibitors[58–60]. Since advanced-cancer is a powerful risk-factor for VTE[49], the most plausible explanation for the association we observed between high TMB and lower rates of VTE, is that it is mediated by tumour stage, as patients with high TMB may have been less likely to develop metastatic disease following study entry.

Thawani and colleagues have previously reported on TMB and VTE risk in a smaller cohort of 1211 patients[61]. In contrast, they did not find an association between TMB and VTE, however it is unclear whether any patients in their study had a TMB≥20. Their analysis may therefore have been underpowered to detect the non-linear association we observed and may not be directly comparable.

We also observed associations between SBS8 and SBS19 and VTE risk. Since the aetiology of both signatures remains unknown[20], biological interpretation of this result is challenging. Although the frequency of SBS8 was high across all cancer groups, SBS19 was predominantly found in haematological malignancies and present in only 5.5% of cancers overall. Our results should therefore be interpreted with caution.

### 4.1 Study limitations

Although most participants recruited to the 100,000 Genomes Project cancer programme had a new cancer diagnosis, 15% of the cohort were enrolled more than 6 months after their initial cancer diagnosis and 167 recruited participants who experienced a VTE in the period between cancer diagnosis and tumour biopsy were excluded from our analysis. Since cancer-associated VTE has a substantial mortality rate[3], we may have under-estimated the VTE incidence associated with some genes, due to survival bias in the recruited cohort.

We note that some rarer tumours, such as sarcoma, were frequent in this cohort, while other tumours, such as prostate cancer were relatively under-represented. Furthermore, only 35% of participants had advanced (stage 3-4) cancer, which potentially reflects a degree of selection bias in the study population. This may limit the generalizability of our findings to an unselected pan-cancer population.

We used electronic health data to identify VTE diagnoses. However, linkage to NHS primary care records was not available. This may have resulted in outcome misclassification for people who were diagnosed with VTE in the community setting alone. Patients with asymptomatic/subclinical VTE who remained undiagnosed would also not have been identified.

We lacked data on several important clinical variables which are likely to modify VTE risk in cancer, including body mass index, blood count parameters and prescriptions of prophylactic anticoagulation[5,6]. Staging data was also missing for 20% of the cohort. This limits our ability to determine whether the VTE risks associated with somatic mutations are independent of other established clinical risks for VTE. Further prospectively designed trials would be helpful to evaluate whether somatic mutation status provides additional clinical utility beyond current VTE risk stratification tools.

Three previous studies have evaluated associations of somatic mutations with VTE in a pan-cancer setting: the largest study[14] analysed 53 cancer-associated genes in 11,695 patients and reported that somatic mutations in *CDKN2B, CTNNB1, KEAP1, KRAS, MET and STK11* were associated with increased rates of VTE, while *SETD2* was associated with lower rates of VTE. Subsequently, two studies[15,16] examining the value of circulating tumour DNA for VTE prediction in smaller pan-cancer cohorts reported that somatic mutations in *TP53*[15]*, KRAS*[15,16]*, EGFR*[15,16]*, PIK3CA*[15]*, CDKN2A*[15,16], *PTEN*[16] and *NF1*[16] were associated with VTE. Several other studies have also previously reported associations between *IDH1* mutations and VTE in glioblastoma, and *ALK* and *ROS1* mutations and VTE in non-small cell lung cancer, as reviewed recently by Lanting *et al*[62].

Of these previously reported associations, we only replicated three loci with confidence (*TP53, KRAS* and *CDKN2A*, all *P*< 0.001). The direction of effect for *EGFR, NF1, KEAP1* and *SETD2* was the same for our analyses compared to previous reports but with wide confidence intervals which encompassed a null effect **(Supplementary Table 5**). We did not observe any pan-cancer associations of *MET*, *CTNNB1, PTEN, STK11, ROS1* and *IDH1* with VTE. *CDKN2B* and *PIK2CA* were not analysed in our study.

In common with the above studies, we collapsed putative pathogenic somatic mutations in each gene into a binary category (mutated vs wildtype), which potentially reduces the power to detect consistent associations, as variants in the same gene which have conflicting effects on transcript abundance or function will likely oppose each other in a regression model. Variation in somatic mutation frequencies and clinical characteristics in this cohort compared to previous studies may also explain some differences in results, since the effect of somatic mutations on VTE risk may vary substantially between different tumour types. Low rates of replication across the different studies may also be attributable to residual confounding from tumour subtype, stage and treatment effects, which are likely to have affected results from all observational studies including this one.

## 5. Conclusion

These findings support the hypothesis that tumour somatic mutation profile influences risk of VTE. However, the mechanisms underlying many of these associations remain unclear, since somatic mutations may proxy clinical tumour characteristics or treatment regimens which contribute to VTE risk. Future work, including meta-analyses of existing results, additional studies of observational cohorts with comprehensive clinical data and mechanistic experimental approaches, would be helpful to triangulate evidence for genetic loci which are implicated in cancer-associated VTE, and to inform future trials investigating the utility of including somatic mutation profile in risk-prediction for cancer-associated VTE.

## Ethics statement

This study was approved by the Genomics England research network and access to the data adhered to Genomics England research governance agreements. All participants agreed to participate in the 100,000 Genomes Project and provided written informed consent. The 100,000 Genomes Project is part of the National Genomics Research Library, approved by the East of England - Cambridge South Research Ethics Committee (14/EE/1112).

## Author Contributions

NC, AM, SW and PH planned the study; MW provided advice on study design and statistical analyses; NC analysed the data and drafted the manuscript; RW, CT, SW and PH were involved in data interpretation. All authors contributed to critical review of the manuscript and approved the final draft.

## Supporting information

STROBE_checklist

## Acknowledgements

This research was made possible through access to data in the National Genomic Research Library, which is managed by Genomics England Limited (a wholly owned company of the Department of Health and Social Care). The National Genomic Research Library holds data provided by patients and collected by the NHS as part of their care and data collected as part of their participation in research. The National Genomic Research Library is funded by the National Institute for Health Research and NHS England.

The Wellcome Trust, Cancer Research UK and the Medical Research Council have also funded research infrastructure. We are very grateful to all the patients who participated in this resource and their families, as well as the team at Genomics England, who supported many aspects of this analysis.

## Data availability

The de-identified patient data used for this analysis can be accessed via the Genomics England Research Environment subject to a collaborative agreement that adheres to patient-led governance. For more information about accessing the data, contact or access the relevant information on the Genomics England website: https://www.genomicsengland.co.uk/research.

Statistical analyses were carried out in R version 4.2.1 using standard software packages including tidyverse, survival and cmprsk[63–65].

## References

1 Cohen AT, Katholing A, Rietbrock S, Bamber L, Martinez C. Epidemiology of first and recurrent venous thromboembolism in patients with active cancer. A population-based cohort study. Thromb Haemost 2017; 117: 57–65.

2 Harry J, Bucciol R, Finnigan D, Hashem H, Araki A, Othman M. The incidence of venous thromboembolism by type of solid cancer worldwide: A systematic review. Cancer Epidemiol 2025; 95: 102764.

3 Sørensen HT, Pedersen L, van Es N, Büller HR, Horváth-Puhó E. Impact of venous thromboembolism on the mortality in patients with cancer: a population-based cohort study. Lancet Reg Health Eur 2023; 34: 100739.

4 Rutjes AW, Porreca E, Candeloro M, Valeriani E, Di Nisio M. Primary prophylaxis for venous thromboembolism in ambulatory cancer patients receiving chemotherapy. Cochrane Database Syst Rev. 2020. p. CD008500.

5 Vladic N, Englisch C, Ay C, Pabinger I. Risk assessment and prevention of cancer-associated venous thromboembolism in ambulatory patients with solid malignancies. Res Pract Thromb Haemost 2025; 9: 102664.

6 Khorana AA, Kuderer NM, Culakova E, Lyman GH, Francis CW. Development and validation of a predictive model for chemotherapy-associated thrombosis. Blood. 2008. p. 4902–7.

7 Mulder FI, Candeloro M, Kamphuisen PW, Di Nisio M, Bossuyt PM, Guman N, Smit K, Büller HR, van Es N, CAT-prediction collaborators. The Khorana score for prediction of venous thromboembolism in cancer patients: a systematic review and meta-analysis. Haematologica 2019; 104: 1277–87.

8 van Es N, Di Nisio M, Cesarman G, Kleinjan A, Otten HM, Mahé I, Wilts IT, Twint DC, Porreca E, Arrieta O, Stépanian A, Smit K, De Tursi M, Bleker SM, Bossuyt PM, Nieuwland R, Kamphuisen PW, Büller HR. Comparison of risk prediction scores for venous thromboembolism in cancer patients: a prospective cohort study. Haematologica. 2017. p. 1494–501.

9 Walker AJ, Card TR, West J, Crooks C, Grainge MJ. Incidence of venous thromboembolism in patients with cancer – A cohort study using linked United Kingdom databases. European Journal of Cancer 2013; 49: 1404–13.

10 Lucotti S, Muschel RJ. Platelets and Metastasis: New Implications of an Old Interplay. Front Oncol 2020; 10: 1350.

11 Palumbo JS, Talmage KE, Massari JV, La Jeunesse CM, Flick MJ, Kombrinck KW, Hu Z, Barney KA, Degen JL. Tumor cell-associated tissue factor and circulating hemostatic factors cooperate to increase metastatic potential through natural killer cell-dependent and-independent mechanisms. Blood 2007; 110: 133–41.

12 Riedl J, Preusser M, Nazari PMS, Posch F, Panzer S, Marosi C, Birner P, Thaler J, Brostjan C, Lötsch D, Berger W, Hainfellner JA, Pabinger I, Ay C. Podoplanin expression in primary brain tumors induces platelet aggregation and increases risk of venous thromboembolism. Blood 2017; 129: 1831–9.

13 Sosinsky A, Ambrose J, Cross W, Turnbull C, Henderson S, Jones L, Hamblin A, Arumugam P, Chan G, Chubb D, Noyvert B, Mitchell J, Walker S, Bowman K, Pasko D, Buongermino Pereira M, Volkova N, Rueda-Martin A, Perez-Gil D, Lopez J, et al. Insights for precision oncology from the integration of genomic and clinical data of 13,880 tumors from the 100,000 Genomes Cancer Programme. Nat Med 2024; 30: 279–89.

14 Dunbar A, Bolton KL, Devlin SM, Sanchez-Vega F, Gao J, Mones JV, Wills J, Kelly D, Farina M, Cordner KB, Park Y, Kishore S, Juluru K, Iyengar NM, Levine RL, Zehir A, Park W, Khorana AA, Soff GA, Mantha S. Genomic profiling identifies somatic mutations predicting thromboembolic risk in patients with solid tumors. Blood 2021; 137: 2103–13.

15 Jee J, Brannon AR, Singh R, Derkach A, Fong C, Lee A, Gray L, Pichotta K, Luthra A, Diosdado M, Haque M, Guo J, Hernandez J, Garg K, Wilhelm C, Arcila ME, Pavlakis N, Clarke S, Shah SP, Razavi P, et al. DNA liquid biopsy-based prediction of cancer-associated venous thromboembolism. Nat Med Nature Publishing Group; 2024; : 1–9.

16 Ma S, Jiang JY, Kim R bum, Chiang E, Theng Tiong JW, Ryu J, Guffey D, Bandyo R, Dowst H, Swinnerton KN, Fillmore NR, La J, Li A. Circulating tumor DNA predicts venous thromboembolism in patients with cancers. Journal of Thrombosis and Haemostasis 2024; .

17 Genomics England. The National Genomics Research Library v5.1. 2020.

18 World Health Organization. International statistical classification of diseases and related health problems (11th ed.). 2019.

19 Bleda M, Tarraga J, de Maria A, Salavert F, Garcia-Alonso L, Celma M, Martin A, Dopazo J, Medina I. CellBase, a comprehensive collection of RESTful web services for retrieving relevant biological information from heterogeneous sources. Nucleic Acids Research 2012; 40: W609– 14.

20 Sondka Z, Dhir NB, Carvalho-Silva D, Jupe S, Madhumita, McLaren K, Starkey M, Ward S, Wilding J, Ahmed M, Argasinska J, Beare D, Chawla MS, Duke S, Fasanella I, Neogi AG, Haller S, Hetenyi B, Hodges L, Holmes A, et al. COSMIC: a curated database of somatic variants and clinical data for cancer. Nucleic Acids Research 2024; 52: D1210–7.

21 Landrum MJ, Lee JM, Benson M, Brown GR, Chao C, Chitipiralla S, Gu B, Hart J, Hoffman D, Jang W, Karapetyan K, Katz K, Liu C, Maddipatla Z, Malheiro A, McDaniel K, Ovetsky M, Riley G, Zhou G, Holmes JB, et al. ClinVar: improving access to variant interpretations and supporting evidence. Nucleic Acids Res 2018; 46: D1062–7.

22 Cornish N, Westbury SK, Warkentin MT, Thirlwell C, Mumford AD, Haycock PC. Association between tumour somatic mutations and venous thromboembolism in the 100,000 Genomes Project cancer cohort: a study protocol. Wellcome Open Res 2024; 9: 640.

23 Martin FJ, Amode MR, Aneja A, Austine-Orimoloye O, Azov AG, Barnes I, Becker A, Bennett R, Berry A, Bhai J, Bhurji SK, Bignell A, Boddu S, Branco Lins PR, Brooks L, Ramaraju SB, Charkhchi M, Cockburn A, Da Rin Fiorretto L, Davidson C, et al. Ensembl 2023. Nucleic Acids Res 2023; 51: D933–41.

24 Klarin D, Busenkell E, Judy R, Lynch J, Levin M, Haessler J, Aragam K, Chaffin M, Haas M, Lindström S, Assimes TL, Huang J, Min Lee K, Shao Q, Huffman JE, Kabrhel C, Huang Y, Sun YV, Vujkovic M, Saleheen D, et al. Genome-wide association analysis of venous thromboembolism identifies new risk loci and genetic overlap with arterial vascular disease. Nat Genet. 2019. p. 1574–9.

25 Lindström S, Wang L, Smith EN, Gordon W, van Hylckama Vlieg A, de Andrade M, Brody JA, Pattee JW, Haessler J, Brumpton BM, Chasman DI, Suchon P, Chen MH, Turman C, Germain M, Wiggins KL, MacDonald J, Braekkan SK, Armasu SM, Pankratz N, et al. Genomic and transcriptomic association studies identify 16 novel susceptibility loci for venous thromboembolism. Blood. 2019. p. 1645–57.

26 Thibord F, Klarin D, Brody JA, Chen M-H, Levin MG, Chasman DI, Goode EL, Hveem K, Teder-Laving M, Martinez-Perez A, Aïssi D, Daian-Bacq D, Ito K, Natarajan P, Lutsey PL, Nadkarni GN, de Vries PS, Cuellar-Partida G, Wolford BN, Pattee JW, et al. Cross-Ancestry Investigation of Venous Thromboembolism Genomic Predictors. Circulation American Heart Association; 2022; 0: 10.1161/CIRCULATIONAHA.122.059675.

27 Ghouse J, Tragante V, Ahlberg G, Rand SA, Jespersen JB, Leinøe EB, Vissing CR, Trudsø L, Jonsdottir I, Banasik K, Brunak S, Ostrowski SR, Pedersen OB, Sørensen E, Erikstrup C, Bruun MT, Nielsen KR, Køber L, Christensen AH, Iversen K, et al. Genome-wide meta-analysis identifies 93 risk loci and enables risk prediction equivalent to monogenic forms of venous thromboembolism. Nat Genet Nature Publishing Group; 2023; : 1–11.

28 He X-Y, Wu B-S, Yang L, Guo Y, Deng Y-T, Li Z-Y, Fei C-J, Liu W-S, Ge Y-J, Kang J, Feng J, Cheng W, Dong Q, Yu J-T. Genetic associations of protein-coding variants in venous thromboembolism. Nat Commun 2024; 15: 2819.

29 Walker DA, Smith TJ. Logistic Regression Under Sparse Data Conditions. J Mod Appl Stat Methods 2020; 18: 2–18.

30 Alexandrov LB, Nik-Zainal S, Wedge DC, Campbell PJ, Stratton MR. Deciphering signatures of mutational processes operative in human cancer. Cell Rep 2013; 3: 246–59.

31 Guman NAM, Mulder FI, Ferwerda B, Zwinderman AH, Kamphuisen PW, Büller HR, van Es N. Polygenic risk scores for prediction of cancer-associated venous thromboembolism in the UK Biobank cohort study. J Thromb Haemost 2023; : S1538-7836(23)00571–8.

32 Linear scoring - PLINK 2.0.

33 Choi SW, Mak TS, O’Reilly PF. Tutorial: a guide to performing polygenic risk score analyses. Nat Protoc. 2020. p. 2759–72.

34 Lau B, Cole SR, Gange SJ. Competing Risk Regression Models for Epidemiologic Data. American Journal of Epidemiology 2009; 170: 244–56.

35 Falanga A, Ay C, Di Nisio M, Gerotziafas G, Langer F, Lecumberri R, Mandala M, Maraveyas A, Pabinger I, Jara-Palomares L, Sinn M, Syrigos K, Young A, Jordan K. Venous thromboembolism in cancer patients: ESMO Clinical Practice Guideline†. Annals of Oncology 2023; .

36 Alikhan R, Gomez K, Maraveyas A, Noble S, Young A, Thomas M, British Society for Haematology. Cancer-associated venous thrombosis in adults (second edition): A British Society for Haematology Guideline. Br J Haematol 2024; .

37 Martin KA, Cameron KA, Lyleroehr MJ, Linder JA, O’Brien M, Hirschhorn LR. Venous thromboembolism prevention in cancer care: implementation strategies to address underuse. Res Pract Thromb Haemost 2023; 7: 102173.

38 Alexandrov LB, Nik-Zainal S, Wedge DC, Aparicio SAJR, Behjati S, Biankin AV, Bignell GR, Bolli N, Borg A, Børresen-Dale A-L, Boyault S, Burkhardt B, Butler AP, Caldas C, Davies HR, Desmedt C, Eils R, Eyfjörd JE, Foekens JA, Greaves M, et al. Signatures of mutational processes in human cancer. Nature Nature Publishing Group; 2013; 500: 415–21.

39 Alexandrov LB, Jones PH, Wedge DC, Sale JE, Campbell PJ, Nik-Zainal S, Stratton MR. Clock-like mutational processes in human somatic cells. Nat Genet Nature Publishing Group; 2015; 47: 1402–7.

40 Abufarhaneh M, Pandya RK, Alkhaja A, Iansavichene A, Welch S, Lazo-Langner A. Association between genetic mutations and risk of venous thromboembolism in patients with solid tumor malignancies: A systematic review and meta-analysis. Thrombosis Research 2022; 213: 47–56.

41 Yu JL, May L, Lhotak V, Shahrzad S, Shirasawa S, Weitz JI, Coomber BL, Mackman N, Rak JW. Oncogenic events regulate tissue factor expression in colorectal cancer cells: implications for tumor progression and angiogenesis. Blood 2005; 105: 1734–41.

42 Mukhopadhyay S, Antalis TM, Nguyen KP, Hoofnagle MH, Sarkar R. Myeloid p53 regulates macrophage polarization and venous thrombus resolution by inflammatory vascular remodeling in mice. Blood 2017; 129: 3245–55.

43 Witkiewicz AK, Knudsen KE, Dicker AP, Knudsen ES. The meaning of p16ink4a expression in tumors. Cell Cycle 2011; 10: 2497–503.

44 Buss H, Handschick K, Jurrmann N, Pekkonen P, Beuerlein K, Müller H, Wait R, Saklatvala J, Ojala PM, Schmitz ML, Naumann M, Kracht M. Cyclin-dependent kinase 6 phosphorylates NF-κB P65 at serine 536 and contributes to the regulation of inflammatory gene expression. PLoS One 2012; 7: e51847.

45 Amulic B, Knackstedt SL, Abu Abed U, Deigendesch N, Harbort CJ, Caffrey BE, Brinkmann V, Heppner FL, Hinds PW, Zychlinsky A. Cell-Cycle Proteins Control Production of Neutrophil Extracellular Traps. Developmental Cell 2017; 43: 449–462.e5.

46 Wang W, Oh S, Koester M, Abramowicz S, Wang N, Tall AR, Welch CL. Enhanced Megakaryopoiesis and Platelet Activity in Hypercholesterolemic, B6-Ldlr−/−, Cdkn2a-Deficient Mice. Circ Cardiovasc Genet 2016; 9: 213–22.

47 Kuo C-L, Murphy AJ, Sayers S, Li R, Yvan-Charvet L, Davis JZ, Krishnamurthy J, Liu Y, Puig O, Sharpless NE, Tall AR, Welch CL. Cdkn2a Is an Atherosclerosis Modifier Locus That Regulates Monocyte/Macrophage Proliferation. *Arteriosclerosis*, Thrombosis, and Vascular Biology American Heart Association; 2011; 31: 2483–92.

48 Hannou SA, Wouters K, Paumelle R, Staels B. Functional genomics of the CDKN2A/B locus in cardiovascular and metabolic disease: what have we learned from GWASs? Trends in Endocrinology & Metabolism 2015; 26: 176–84.

49 Noble S, Pasi J. Epidemiology and pathophysiology of cancer-associated thrombosis. Br J Cancer Nature Publishing Group; 2010; 102: S2–9.

50 Guntupalli SR, Spinosa D, Wethington S, Eskander R, Khorana AA. Prevention of venous thromboembolism in patients with cancer. BMJ British Medical Journal Publishing Group; 2023; 381: e072715.

51 Delmaghani S, El-Amraoui A. The genetic and phenotypic landscapes of Usher syndrome: from disease mechanisms to a new classification. Hum Genet 2022; 141: 709–35.

52 Moreno CS, Winham CL, Alemozaffar M, Klein ER, Lawal IO, Abiodun-Ojo OA, Patil D, Barwick BG, Huang Y, Schuster DM, Sanda MG, Osunkoya AO. Integrated Genomic Analysis of Primary Prostate Tumor Foci and Corresponding Lymph Node Metastases Identifies Mutations and Pathways Associated with Metastasis. Cancers (Basel) 2023; 15: 5671.

53 Xu S, Moss TJ, Laura Rubin M, Ning J, Eterovic K, Yu H, Jia R, Fan X, Tetzlaff MT, Esmaeli B. Whole-exome sequencing for ocular adnexal sebaceous carcinoma suggests PCDH15 as a novel mutation associated with metastasis. Mod Pathol 2020; 33: 1256–63.

54 Han M, Wang S, Fritah S, Wang X, Zhou W, Yang N, Ni S, Huang B, Chen A, Li G, Miletic H, Thorsen F, Bjerkvig R, Li X, Wang J. Interfering with long non-coding RNA MIR22HG processing inhibits glioblastoma progression through suppression of Wnt/β-catenin signalling. Brain 2020; 143: 512–30.

55 Gil-Bernabé AM, Ferjancic S, Tlalka M, Zhao L, Allen PD, Im JH, Watson K, Hill SA, Amirkhosravi A, Francis JL, Pollard JW, Ruf W, Muschel RJ. Recruitment of monocytes/macrophages by tissue factor-mediated coagulation is essential for metastatic cell survival and premetastatic niche establishment in mice. Blood. 2012. p. 3164–75.

56 Gong T, Hayes VM, Chan EKF. Detection of somatic structural variants from short-read next-generation sequencing data. Brief Bioinform 2020; 22: bbaa056.

57 Yang RK, Alvarez H, Lucas AS, Roy-Chowdhuri S, Rashid A, Chen H, Ballester LY, Sweeney K, Routbort MJ, Patel KP, Luthra R, Medeiros LJ, Toruner GA. Microsatellite instability and high tumor mutational burden detected by next generation sequencing are concordant with loss of mismatch repair proteins by immunohistochemistry. Cancer Genetics 2025; 290–291: 44–50.

58 Manca P, Corti F, Intini R, Mazzoli G, Miceli R, Germani MM, Bergamo F, Ambrosini M, Cristarella E, Cerantola R, Boccaccio C, Ricagno G, Ghelardi F, Randon G, Leoncini G, Milione M, Fassan M, Cremolini C, Lonardi S, Pietrantonio F. Tumour mutational burden as a biomarker in patients with mismatch repair deficient/microsatellite instability-high metastatic colorectal cancer treated with immune checkpoint inhibitors. European Journal of Cancer Elsevier; 2023; 187: 15–24.

59 Antoniotti C, Korn WM, Marmorino F, Rossini D, Lonardi S, Masi G, Randon G, Conca V, Boccaccino A, Tomasello G, Passardi A, Swensen J, Ugolini C, Oberley M, Tamburini E, Casagrande M, Domenyuk V, Fontanini G, Giordano M, Abraham J, et al. Tumour mutational burden, microsatellite instability, and actionable alterations in metastatic colorectal cancer: Next-generation sequencing results of TRIBE2 study. European Journal of Cancer 2021; 155: 73–84.

60 Gandara DR, Agarwal N, Gupta S, Klempner SJ, Andrews MC, Mahipal A, Subbiah V, Eskander RN, Carbone DP, Riess JW, Sammons S, Snider J, Bouzit L, Cho-Phan C, Price M, Li G, Quintanilha JCF, Huang RSP, Ross JS, Fabrizio D, et al. Tumor mutational burden and survival on immune checkpoint inhibition in >8000 patients across 24 cancer types. J Immunother Cancer 2025; 13: e010311.

61 Thawani R, Kartika T, Elstrott B, Batiuk E, Tao D, Gowda S, Chen L, Lavasseur C, Tun N, Taflin NF, Shatzel J. Association of PD-L1 expression, tumor mutational burden and immunotherapy with venous thrombosis in patients with solid organ malignancies. Thromb Res 2022; 217: 12–4.

62 Lanting V, Oskam M, Wilmink H, Kamphuisen PW, van Es N. The role of germline and somatic mutations in predicting cancer-associated thrombosis: a narrative review. Curr Opin Hematol 2025; .

63 Wickham H, Averick M, Bryan J, Chang W, McGowan LD, François R, Grolemund G, Hayes A, Henry L, Hester J, Kuhn M, Pedersen TL, Miller E, Bache SM, Müller K, Ooms J, Robinson D, Seidel DP, Spinu V, Takahashi K, et al. Welcome to the Tidyverse. Journal of Open Source Software 2019; 4: 1686.

64 Therneau TM, until 2009) TL (original S->R port and R maintainer, Elizabeth A, Cynthia C. survival: Survival Analysis. 2024.

65 Gray B. cmprsk: Subdistribution Analysis of Competing Risks. 2024.

