## Supplementary Methods for "Associations of tumour somatic mutations with cancer-associated venous thromboembolism"

### Whole genome sequencing and variant calling

Tumour DNA was obtained primarily from fresh-frozen histology specimens (and occasionally from formalin-fixed paraffin-embedded samples), while germline DNA was obtained from blood or saliva. DNA extraction was performed in local laboratories linked to the regional NHS Genomic Medicine Centres, as described in the Genomics England[1] (GEL) Sample Handling Guidance (v.4.0) available at [Document library | Genomics England].

Samples were prepared with an Illumina TruSeq PCR-free library preparation kit, providing sufficient DNA was available; otherwise, PCR-based library preparation was used for a minority of samples. DNA sequencing was performed centrally on an Illumina HiSeq platform to an average coverage of 30x (germline DNA) and 100x (tumour DNA). Sequence data was processed using the Illumina North Star (version 2.6.53.23) pipeline with read alignment against the human reference genome GRCh38-Decoy+EBV using ISAAC (version iSAAC-03.16.02.19).

Germline SNVs were called by Strelka (v.2.4.7) [https://github.com/Illumina/strelka] and genetic principal components (PCs) were calculated by GEL bioinformaticians using GCTA[2] version 1.92.4. PCs were derived from 63,523 high confidence LD-pruned bi-allelic autosomal non-palindromic SNPs with a minor allele frequency >5% meeting quality thresholds (missingness <1%, genotype quality >= 30, median read-depth >= 30, Hardy-Weinberg equilibrium p value > $1 \times 10^{-5}$). Genetic ancestry (categorized according to the five broad-continental superpopulations) was inferred by GEL by projecting PCs for each participant onto a PC matrix from the 1000 genomes project phase 3 (1KGP3)[3].

Somatic small variants, including single nucleotide variants (SNVs) and indels < 50bp, were identified using Strelka (v2.4.7) Large structural variants (including inversions, translocations and indels >50bp) were called by Manta (v.0.28.09) [https://github.com/Illumina/manta].

Variants which were present in the paired germline sequence were subtracted so that only somatic variants were retained.

In addition to the default filters used by the variant-calling software, we removed the following small variants flagged by GEL[5] as being potentially unreliable: 1) SNVs with a population germline allele frequency >1% in the GEL or gnomAD[4] datasets (these may represent residual germline variants which have been mistakenly called as somatic); 2) recurrent somatic variants occurring in >5% of the cohort (likely technical sequencing artefacts); 3) variants overlapping simple repeats or intersecting reference homopolymers 4) small indels in regions of high sequencing noise (where the proportion of low quality Strelka-filtered base-calls within 50bp of the variant exceeded 10%); 5) variants flagged as systematic sequencing/mapping errors (Fisher's exact test Phred score < 50). 6) variant allele proportion (number of alternate reads divided by total reads) < 0.05.[5]

For the gene-level analyses, we filtered for small somatic variants annotated as 'pathogenic/likely pathogenic' in either COSMIC (v95)[6] or Clinvar (2018)[7] and any variant predicted by Cellbase annotation (v.4.7.1)[8] to result in transcript ablation, frameshift, start or stop codon disruption or splice site disruption as well as in-frame insertions or deletions. Variants >50bp were included in the analysis if they were present in the GEL 'tiered-variant' JSON files, which are produced from an internal pipeline which filters the Manta variant-call files for structural variants predicted to disrupt the canonical transcript of a gene, based on transcript orientation and SV breakpoint.

**WGS quality thresholds for inclusion in study**

- WGS read-mapping quality > 30 across 210 Gb for tumour DNA and 85Gb for germline DNA
- cross-sample contamination <3% for germline DNA (assessed by VerifyBamID [https://github.com/statgen/verifyBamID] or <5% for tumour DNA (assessed by ConPair [https://github.com/nygenome/Conpair]).
- concordant phenotypic and karyotypic sex
- Genetic principal components calculated by GEL

**Deviations from a-priori study protocol**

We published a peer-reviewed study protocol prior to starting the analysis: https://wellcomeopenresearch.org/articles/9-640/v2

Subsequent deviations from the protocol were as follows:

1) The protocol stated that cancer type would be grouped by ICD10 code (for covariate adjustment and tumour-stratified analyses). We instead applied the cancer 'disease type' category recorded by Genomics England (GEL) during recruitment as this aligned with the existing data structure. Comparisons with ICD10 codes recorded in the National cancer registry (NCRAS) record were performed for each participant and participants were excluded where the GEL disease type conflicted with the WHO ICD10 version:2019[9] level 2 mapped code in the NCRAS record (see supplementary table 2).
2) The protocol stated that patients with missing covariate information would be excluded from analyses. However, 20% of the cohort did not have cancer stage recorded and missing staging data was likely biased towards specific tumour types (including haematological cancers). Therefore, ‘unavailable’ stage was included as a distinct category for the staging covariate.
3) The protocol stated that analyses would be restricted to genetic variables where at least 5 VTE events occurred in each category (i.e. wildtype vs mutated categories). For the primary analyses, we limited analyses to variables with at least 10 VTE events per category. This was done to reduce the risk of unreliable results from sparse data. For sensitivity analyses (including tumour-stratified and ancestry-stratified analyses), no lower limit was applied but results were not reported if the regression model failed to converge.
4) The protocol stated that we would report the hazard ratio (HR) for VTE per standard deviation increase in tumour mutational burden (TMB). However, on evaluating statistical assumptions underlying this model we found the association between TMB and VTE appeared non-linear, therefore we report the HR for VTE in each TMB category: 0-4 mutations/Mb (reference category), 5-9 mutations/Mb, 10-19 mutations/Mb, 20 mutations/Mb.

5) We originally planned a sensitivity analysis where patients with a delay >180 days between cancer diagnosis and study entry were excluded. We subsequently changed this exclusion threshold to >42 days delay between cancer diagnosis and study entry. This approach was considered preferable to limit bias resulting from misalignment of exposure allocation and study eligibility.

**References**


1. Genomics England. The National Genomics Research Library v5.1 [Internet]. 2020 [cited 2024 Sept 1];Available from: doi:10.6084/m9.figshare.4530893/7. 2020.

2. Yang J, Lee SH, Goddard ME, Visscher PM. GCTA: a tool for genome-wide complex trait analysis. Am J Hum Genet 2011;88(1):76–82.

3. Auton A, Abecasis GR, Altshuler DM, Durbin RM, Abecasis GR, Bentley DR, et al. A global reference for human genetic variation. Nature 2015;526(7571):68–74.

4. Chen S, Francioli LC, Goodrich JK, Collins RL, Kanai M, Wang Q, et al. A genomic mutational constraint map using variation in 76,156 human genomes. Nature 2024;625(7993):92–100.

5. Genomics England. Cancer Analysis Technical Information Document, Genomics England [Internet]. 2019 [cited 2024 Sept 26];Available from: https://www.genomicsengland.co.uk/initiatives/100000-genomes-project/documentation

6. Sondka Z, Dhir NB, Carvalho-Silva D, Jupe S, Madhumita, McLaren K, et al. COSMIC: a curated database of somatic variants and clinical data for cancer. Nucleic Acids Research 2024;52(D1):D1210–7.

7. Landrum MJ, Lee JM, Benson M, Brown GR, Chao C, Chitipiralla S, et al. ClinVar: improving access to variant interpretations and supporting evidence. Nucleic Acids Res 2018;46(D1):D1062–7.

8. Bleda M, Tarraga J, de Maria A, Salavert F, Garcia-Alonso L, Celma M, et al. CellBase, a comprehensive collection of RESTful web services for retrieving relevant biological information from heterogeneous sources. Nucleic Acids Research 2012;40(W1):W609–14.

9. World Health Organization. International statistical classification of diseases and related health problems (11th ed.) [Internet]. 2019 [cited 2024 July 31];Available from: https://icd.who.int/browse10/2019/en#/I80-I89
