## Supplementary Figures for "Associations of tumour somatic mutations with cancer-associated venous thromboembolism"

**CONTENTS**

**Supplementary Figure 1**: Diagram showing the hypothesised relationships between potential confounders (grey) and mediators (green) of the associations between tumour somatic mutation profile and VTE.

**Supplementary Figure 2***:* Instantaneous rate of VTE over study period**.**

**Supplementary Figure 3:** Cumulative incidence estimates for VTE after accounting for death as a competing risk factor, comparing wildtype sequence (blue) vs somatic mutations (red) in A) *TP53*, B) *KRAS*, C) *CDKN2A*, D) *PCDH15*

**Supplementary Figure 4:** Distribution of tumour mutational burden (TMB) by tumour type.

**Supplementary Figure 5**: 'Leave one out' analyses (pan-cancer estimates after sequentially excluding each tumour type) for associations between very high tumour mutational burden (TMB)≥20 mutations/Mb (reference category TMB<20 mutations/Mb) and VTE

**Supplementary Figure 6**: Presence of single base substitution (SBS) mutational signatures (SBS6, SBS8, SBS19 and SBS26) across different tumour types. The plots show the median value and interquartile range (bars) for the proportional contribution of each signature to TMB across each participant's tumour sample. The percentage given above each bar represents the overall proportion of tumour samples where the signature was present at any level.

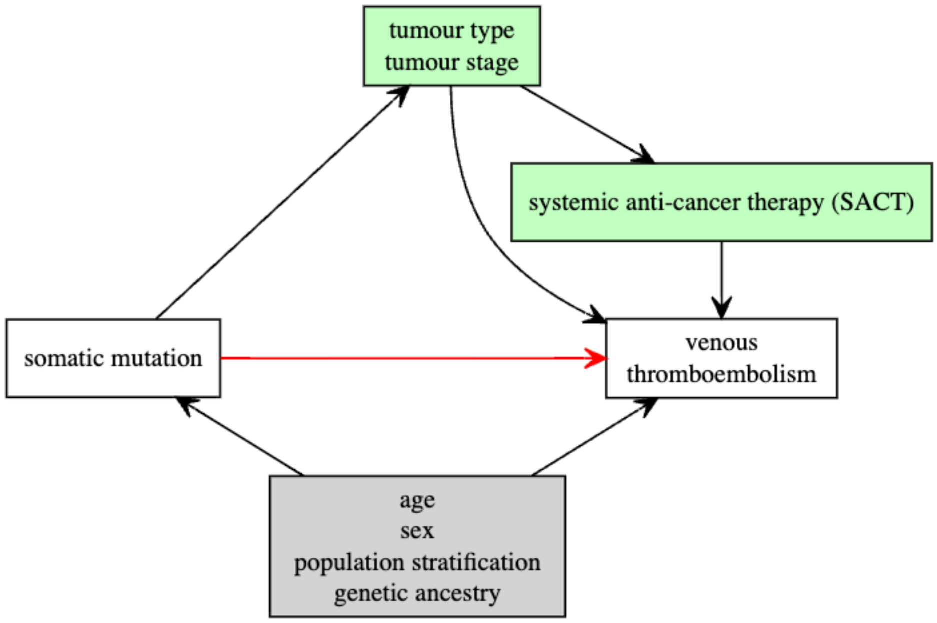


Supplementary Figure 1: Diagram showing the hypothesised relationships between potential confounders (grey) and mediators (green) of the associations between tumour somatic mutation profile and VTE.

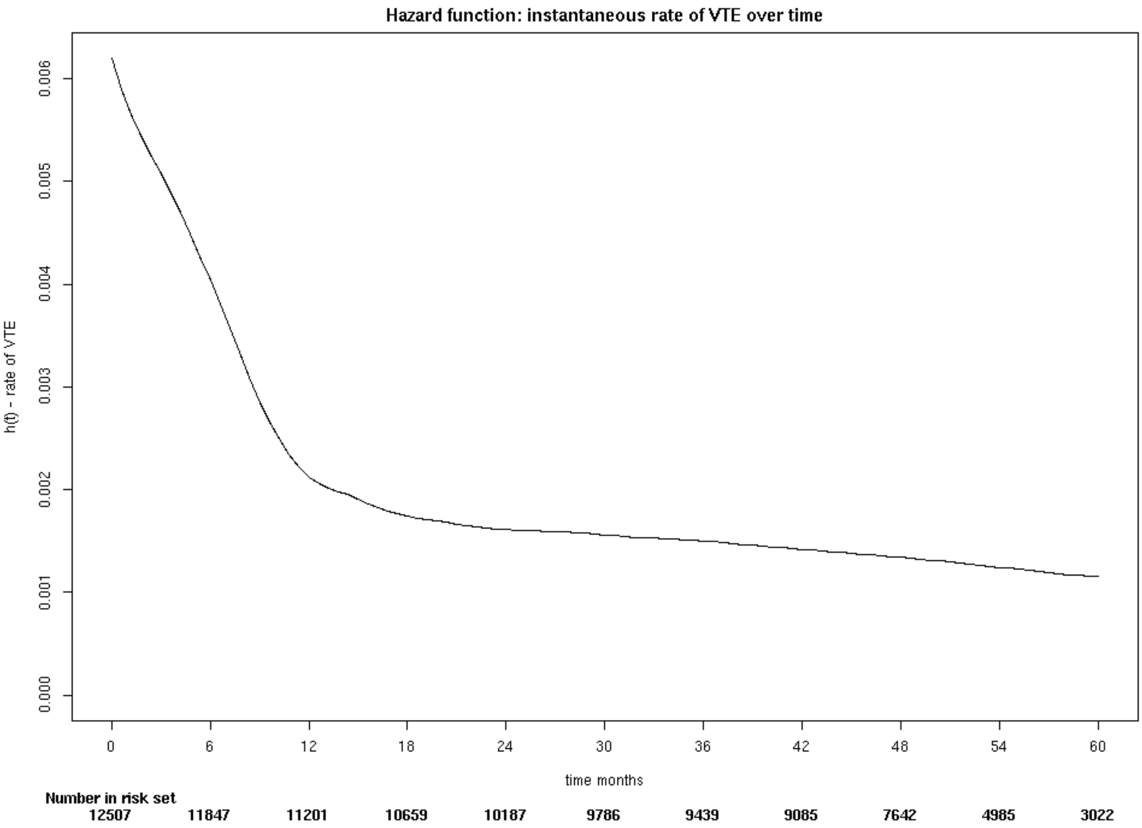


Supplementary Figure 2*:* Instantaneous rate of VTE over study period.

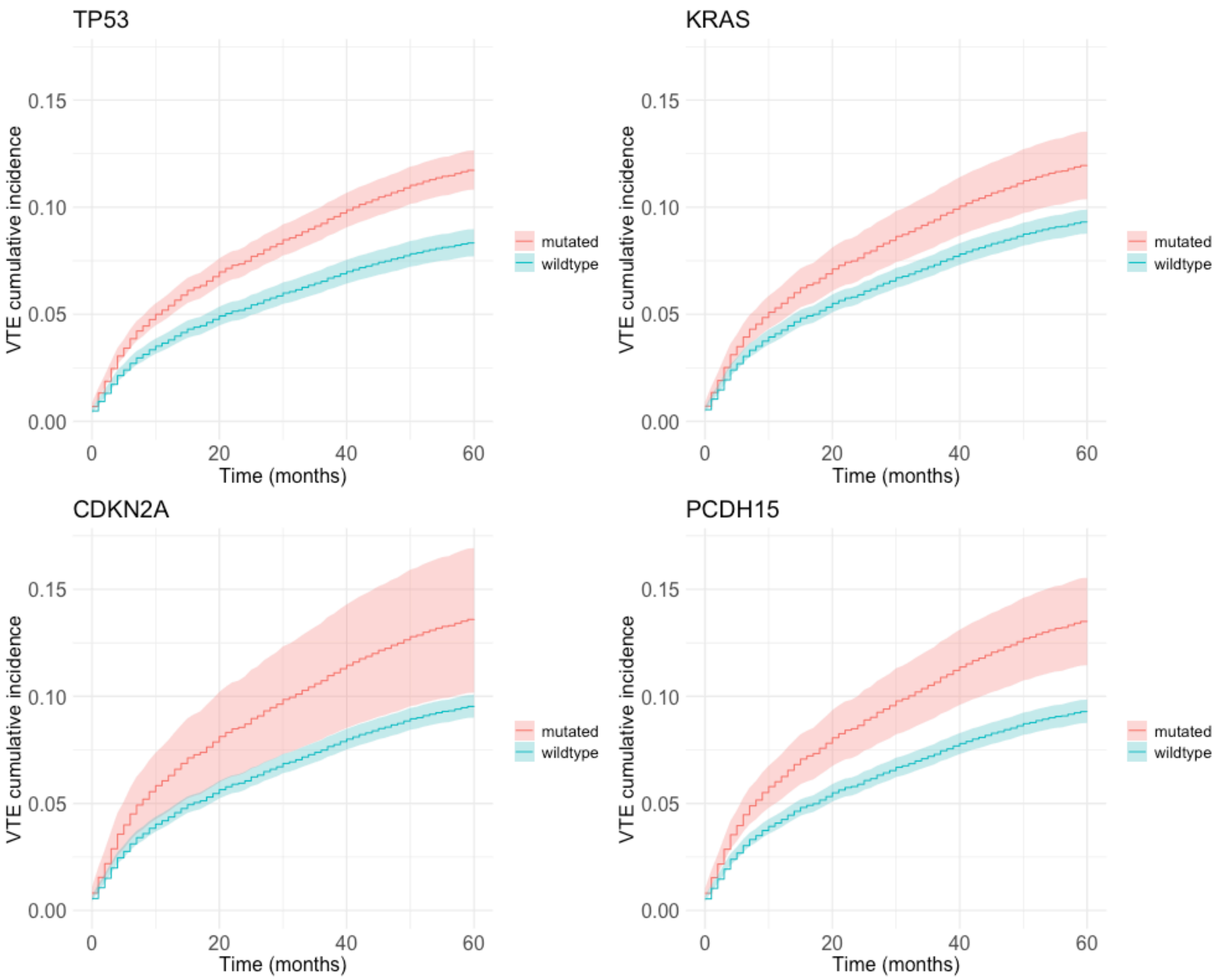


Supplementary Figure 3: Cumulative incidence estimates for VTE after accounting for death as a competing risk factor, comparing wildtype sequence (blue) vs somatic mutations (red) in *TP53*, *KRAS*, *CDKN2A*, *PCDH15*

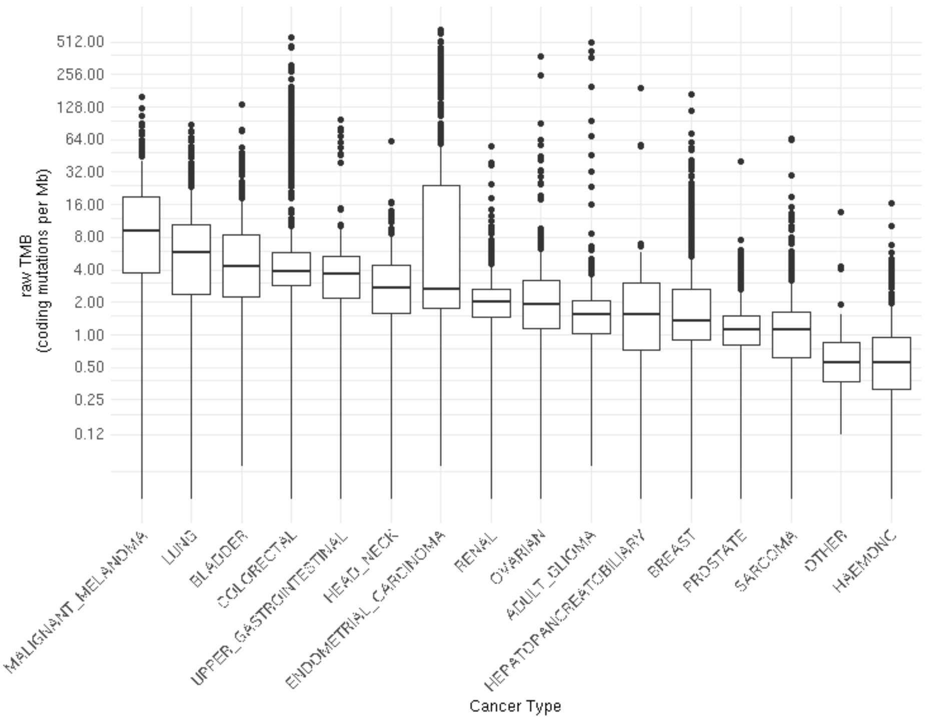


Supplementary Figure 4: TMB distribution categorised by tumour type. TMB distribution was positively skewed for most tumours; the y-axis is plotted on a log2 scale to show the distribution of values more clearly.

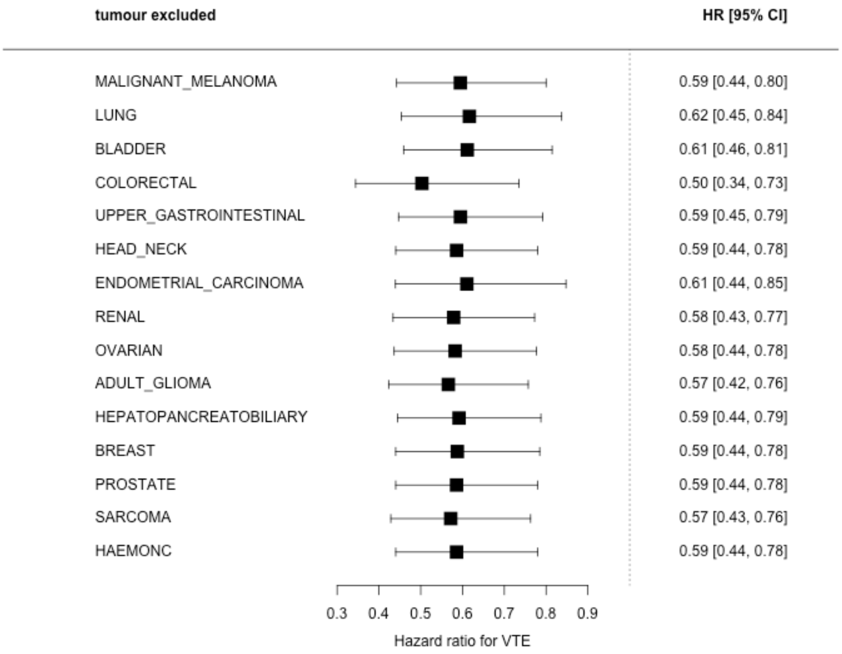


Supplementary Figure 5: ‘Leave one out’ analyses for associations between TMB≥20 mutations/Mb (reference category TMB<20 mutations/Mb) and VTE. The pan-cancer estimates after systematically excluding one cancer type at a time are shown. All analyses adjusted for age, sex, top four genetic principal components, tumour type, stage SACT>6 weeks prior to study entry and SACT during study period.

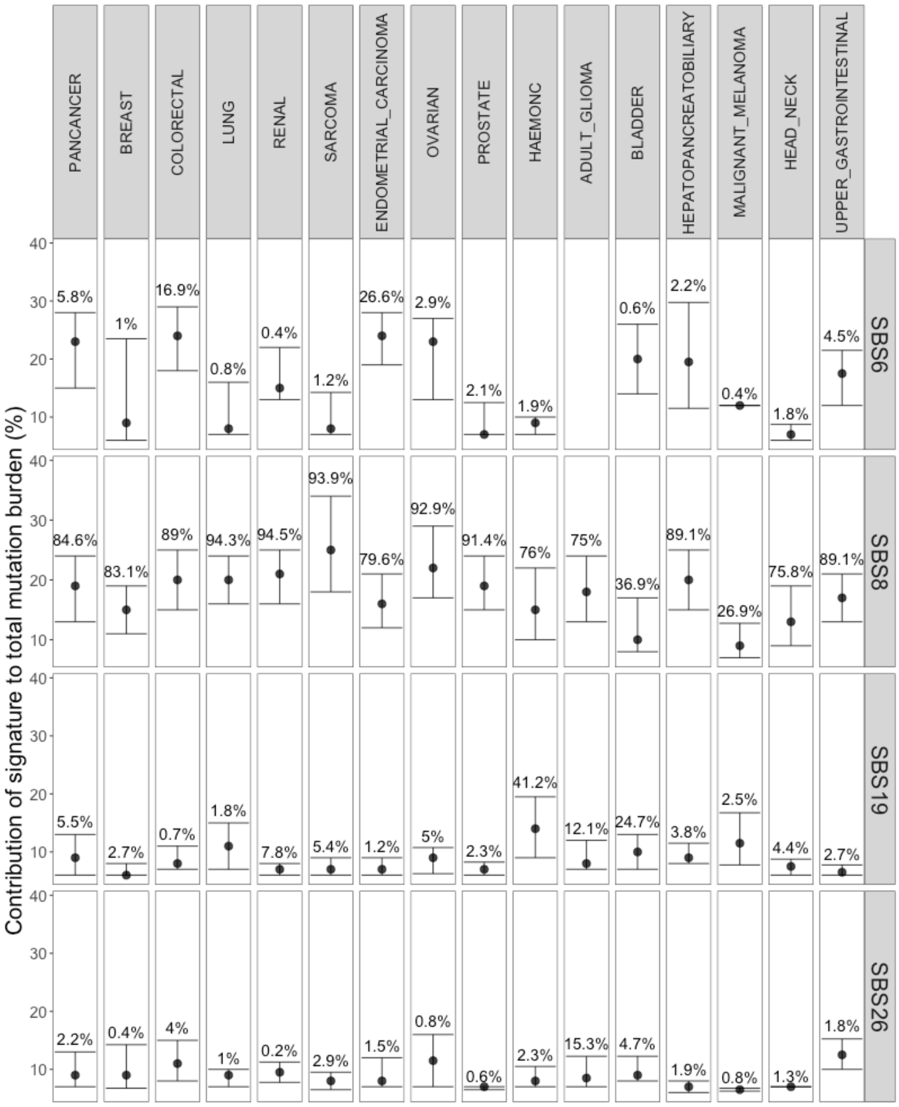


Supplementary Figure 6: Presence of single base substitution (SBS) mutational signatures (SBS6, SBS8, SBS19 and SBS26) across different tumour types. The plots show the median value and interquartile range (bars) for the proportional contribution of each signature to TMB across each participant's tumour sample. The percentage given above each bar represents the overall proportion of tumour samples where the signature was present at any level.
